# Cov-MS: a community-based template assay for clinical MS-based protein detection in SARS-CoV-2 patients

**DOI:** 10.1101/2020.11.18.20231688

**Authors:** B. Van Puyvelde, K. Van Uytfanghe, O. Tytgat, L. Van Oudenhove, R. Gabriels, R. Bouwmeester, S. Daled, T. Van Den Bossche, P. Ramasamy, S. Verhelst, L. De Clerck, L. Corveleyn, N. Debunne, E. Wynendaele, B. De Spiegeleer, P. Judak, K. Roels, L. De Wilde, P. Van Eenoo, T. Reyns, M. Cherlet, E. Dumont, G. Debyser, R. t’Kindt, K. Sandra, S. Gupta, Nicolas Drouin, Amy Harms, Thomas Hankemeier, DJL Jones, P. Gupta, D. Lane, C.S. Lane, S. El Ouadi, JB. Vincendet, N. Morrice, S. Oehrle, N. Tanna, S. Silvester, S. Hannam, F. Sigloch, A. Bhangu-Uhlmann, J. Claereboudt, L. Anderson, M. Razavi, S. Degroeve, L. Cuypers, C. Stove, K. Lagrou, G. Martens, D. Deforce, L. Martens, J.P.C. Vissers, M. Dhaenens

## Abstract

Rising population density and global mobility are among the reasons why pathogens such as SARS-CoV-2, the virus that causes COVID-19, spread so rapidly across the globe. The policy response to such pandemics will always have to include accurate monitoring of the spread, as this provides one of the few alternatives to total lockdown. However, COVID-19 diagnosis is currently performed almost exclusively by Reverse Transcription Polymerase Chain Reaction (RT-PCR). Although this is efficient, automatable and acceptably cheap, reliance on one type of technology comes with serious caveats, as illustrated by recurring reagent and test shortages. We therefore developed an alternative diagnostic test that detects proteolytically digested SARS-CoV-2 proteins using Mass Spectrometry (MS). We established the Cov-MS consortium, consisting of fifteen academic labs and several industrial partners to increase applicability, accessibility, sensitivity and robustness of this kind of SARS-CoV-2 detection. This in turn gave rise to the Cov-MS Digital Incubator that allows other labs to join the effort, navigate and share their optimizations, and translate the assay into their clinic. As this test relies on viral proteins instead of RNA, it provides an orthogonal and complementary approach to RT-PCR, using other reagents that are relatively inexpensive and widely available, as well as orthogonally skilled personnel and different instruments. Data are available via ProteomeXchange with identifier PXD022550.

## Introduction

COVID-19 has disrupted daily life in a substantial part of the world. The virus has forced many countries worldwide to impose lockdowns or take similar wide-ranging measures. To exit such lockdowns, widespread diagnostic capabilities are required to prevent reoccurrence of outbreaks (1). Today, millions of RT-PCR-based tests are performed every day worldwide. Although these are efficient, relatively simple and acceptably cheap, reliance on one type of technology comes with notable disadvantages. Not only does this regularly lead to shortages of the necessary reagents and lack of scalable capacity, it also means that these tests are difficult to validate in terms of sensitivity, false positives and false negative diagnoses (2–4). Additionally, the fact that the presence of nucleic acid alone cannot be used to define viral shedding or infection potential increases the relevance of the detection of other biomolecules (5,6). Therefore, numerous publications aim at expanding the viral targets beyond RNA (7,8,17,9–16). Like many other viruses, SARS-CoV-2 can produce virus-like particles (VLPs) that contain a high concentration of proteins, but no RNA, making these biomolecules very attractive targets for LC-MS based detection and characterization (18,19). In fact, MS-based detection of proteins is directly compatible with conventional clinical small molecule detection setups and is adoptable to new mutations in a matter of hours, a very relevant feature in light of e.g. the recent D614G variant (20). Additionally, the assay is much less prone to contamination than RT-PCR based assays because of the lack of amplification steps. Thus, in contrast with RT-PCR setups, no pre- and post-PCR lab infrastructure needs to be set up, making a roll-out of an MS-based assay relatively easy in comparison. However, when compared to RT-PCR, MS instrumentation and matrix effects from e.g. transport media are more pronounced and no single working protocol can be expected to be easily implementable across all clinical labs.

Therefore, we here describe a community-based effort that has led to a generic and broadly applicable mass spectrometry (MS)-based template assay that allows easy adaptation to the broadly variable testing facility landscape (**Figure 1**). First, we used our high-resolution MS instrument data to create a vendor-independent Skyline document with 17 biomarker peptides from two structural SARS-CoV-2 proteins detected in public data, i.e. Nucleocapsid (NCAP) and Spike (SPIKE2) protein (16,21). We then established a consortium (MS-Cov), consisting of academic and industrial labs, complemented with representation from major MS vendors (Waters Corporation, Sciex, and Thermo Fisher Scientific). All members of this consortium were provided with the Skyline document together with a sample kit containing both pure recombinant NCAP and SPIKE protein as well as a dilution series in a negative patient background. Fifteen labs shared data from four different instrument vendors, which was gathered centrally and used in a vendor-independent freeware (Skyline) to assess overall performance and suitability of the candidate signature peptides (22). Finally, we have established the **Cov-MS Digital Incubator** in the form of a Microsoft Teams environment where open communication and data sharing will help translate this assay into different clinical environments around the globe. **All Supplementary Data is available in the Cov-MS Digital Incubator**. Access requests can be sent to.

We here present a very detailed and fully accessible report of a template assay that will enable labs to assess the feasibility of SARS-CoV-2 MS-based detection for their platform, navigate and share their optimizations and quickly adopt the assay in a common effort to significantly increase testing capacity worldwide in the upcoming months. Shortly, we will become part of COVID-MSC, (23). The LC-MS data generated within the Cov-MS consortium is shared and browsable through Panorama Public (https://panoramaweb.org/CovMS.url) and DDA, predicted and chromatogram libraries as well as the narrow window DIA data have been deposited to the ProteomeXchange Consortium via the PRIDE partner repository (24) with the dataset identifier PXD022550 and 10.6019/PXD022550.

## Results and Discussion

**Figure 1** depicts a schematic overview of the MRM-assay development workflow from discovery to envisioned implementation. Each step is discussed in more detail below in a comprehensive and accessible manner that enables re-analysis of the data online through Panorama Public interface for a broad outreach.

**Figure 1.**
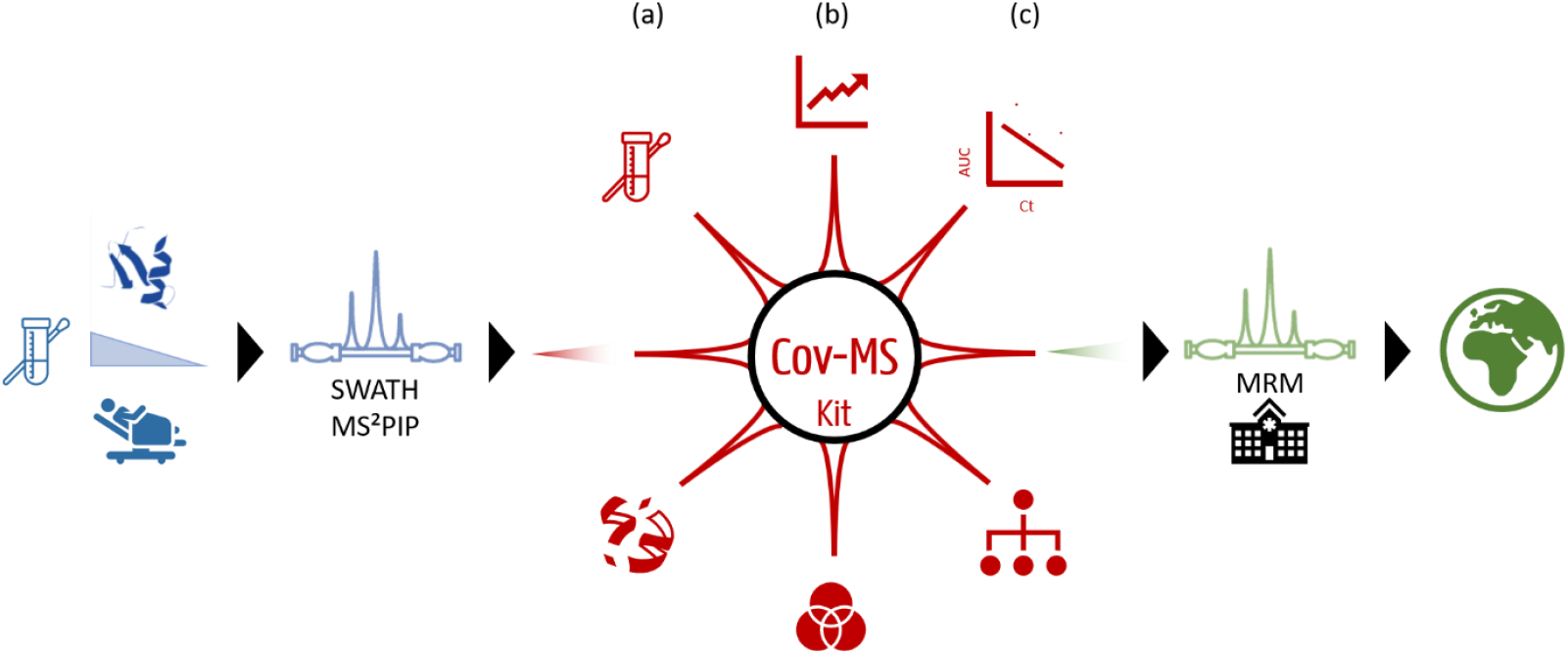
Cov-MS MRM assay development. The SARS-CoV-2 peptide-biomarker discovery workflow **(Blue)** was initiated in mid-March 2020. Using the recombinant NCAP and SPIKE protein (Sino Biological, Beijing, China), combined with twenty nasopharyngeal swabs from patients, we applied our recently published data acquisition and data analysis workflow using machine learning-based spectral predictions (25). With the 17 responsive peptides discovered in this way, a preliminary MRM assay was developed that could classify all twenty blinded patient samples correctly. We then assembled the Cov-MS Consortium **(Red)** comprising of MRM experts from academia and industry, a protein standard company, and a large computational research group. This consortium further optimized sensitivity and robustness using our in-house developed Cov-MS kits. **(a) Optimization of sample preparation** increased digestion efficiency, reduced the time investment down to twenty minutes and assessed alternative transport media and consumables. **(b) Optimization of data acquisition** involved fifteen different labs with different instrumental platforms. These were supported by acquisition specialists from three main instrument vendors. By centralizing all data, instrument-specific peptide targets could be distilled. **(c) Optimization of data analysis** centered around the correlation between MS signal intensity and clinical RT-PCR assay Ct value. Peptides with the highest diagnostic value were selected for each medium through machine learning. Additionally, all peptide biomarker candidates were mapped to the 3D structure of the target proteins and investigated for SARS-CoV-2 strain specificity. To enable future clinical roll out **(Green)**, a heavy QConCAT internal standard was synthesized that enables assessing patient sampling quality, sample preparation efficiency, instrumental robustness and absolute quantification of the viral load. We validated the optimized workflow on 135 patient samples. Finally, we established a Microsoft Teams environment (Cov-MS Digital Incubator) to facilitate global collaboration on the translation of this assay into the clinic.

### Discovery Phase (blue): finding SARS-CoV-2 peptide biomarkers

#### Sample preparation

From the start, sample preparation was designed for typical nasopharyngeal swabs. The aim was to minimize sample consumption, time investment, cost, and reagent use, and to facilitate parallelization. Therefore, immediate acetone precipitation both isolates proteins and inactivates the virus to allow sample preparation in a validated biological safety cabinet (BSC) in Biosafety Level L2 (BSL-2) labs (26). The sample preparation protocol was initially developed on Universal Transport Medium (UTM) swabs from healthy volunteers spiked with recombinant SARS-CoV-2 proteins. It requires only ten minutes of hands-on work and proved applicable down to 1/1000th of the volume for each injection in a conventional UTM sample kit of 3 ml, i.e. 3 µl medium equivalent on column (oc). (**Detailed Methods Section**)

#### Peptide biomarker discovery

The recombinant proteins were analyzed and a spectral library was created for early assay development based on fractionation of the sample combined with data-dependent acquisition (DDA) on high resolution instruments (**Supplementary Data 1a**) (27–29). However, we have recently shown that predicted spectral libraries can be combined with Gas Phase Fractionated (GPF) narrow-window data-independent acquisition (DIA) to generate empirically calibrated chromatogram libraries that are able to detect peptides at lower abundance levels with reduced processing time (30). More importantly, however, narrow window DIA data generates an exhaustive list of detectable pseudo transitions with their respective extracted ion chromatograms (XIC) and interferences (31). This data is akin to parallel reaction monitoring (PRM) data and in its basic form is identical to what is generated on tandem quadrupole instruments operated in MRM acquisition mode. Therefore, creating a targeted method for any tandem quadrupole instrument becomes trivial, i.e. selecting the transitions that are best detected on a given tandem quadrupole MS instrument in a given lab. We therefore opted to share the assay with the community in this form.

First, all tryptic spectra that can be expected from the NCAP_SARS2 and SPIKE_SARS2 proteins were predicted using MS^2^PIP, and their retention times were predicted by DeepLC. From these predictions, a spectral library was created for early assay development (**Supplementary data 1b**) (15,32,33). Using eight GPF runs, a chromatogram library was created from this spectral library (**Supplementary Data 1c**) (34). Next, SARS-CoV-2 peptide signals were extracted from the dilution series that were run by a 64 window SWATH on 75 minutes and 20 minute LC-gradients (**Supplementary Data 2a and 2b**) using a high resolution TripleTOF 6600+ Q-TOF instrument (Sciex, Concord, Canada). We detected seventeen responsive peptides that showed an increase in signal with increased loading (**Figure 2A, Supplementary Data 2c**). At least nine of these have been reported in other studies, confirming that these are indeed interesting targets (17). Notably, one peptide (IGMEVTPSGTWLTYTGAIK) from the recombinant protein was not detected due to a single amino acid substitution in the sequence (G16A). Indeed, Grossegesse M. et al. recently cautioned for the pitfalls of using recombinant proteins for target selection (17). Next, data for the twenty blinded patient samples were acquired on a 75 minute gradient using the 64 window SWATH-mode (**Supplementary Data 3a**). Through manual inspection of the data, we correctly classified 18/20 samples (**Supplementary Data 3b**). The two positive patients that were classified as negative at this point had the lowest viral load according to Real Time Polymerase Chain Reaction (RT-PCR), *i*.*e*. with the highest Ct values of 19 and 20. Notably, the exploratory DDA approach had detected SARS-CoV-2 peptides in only two of these twenty patients (Supplementary Data 3c).

**Figure 2:**
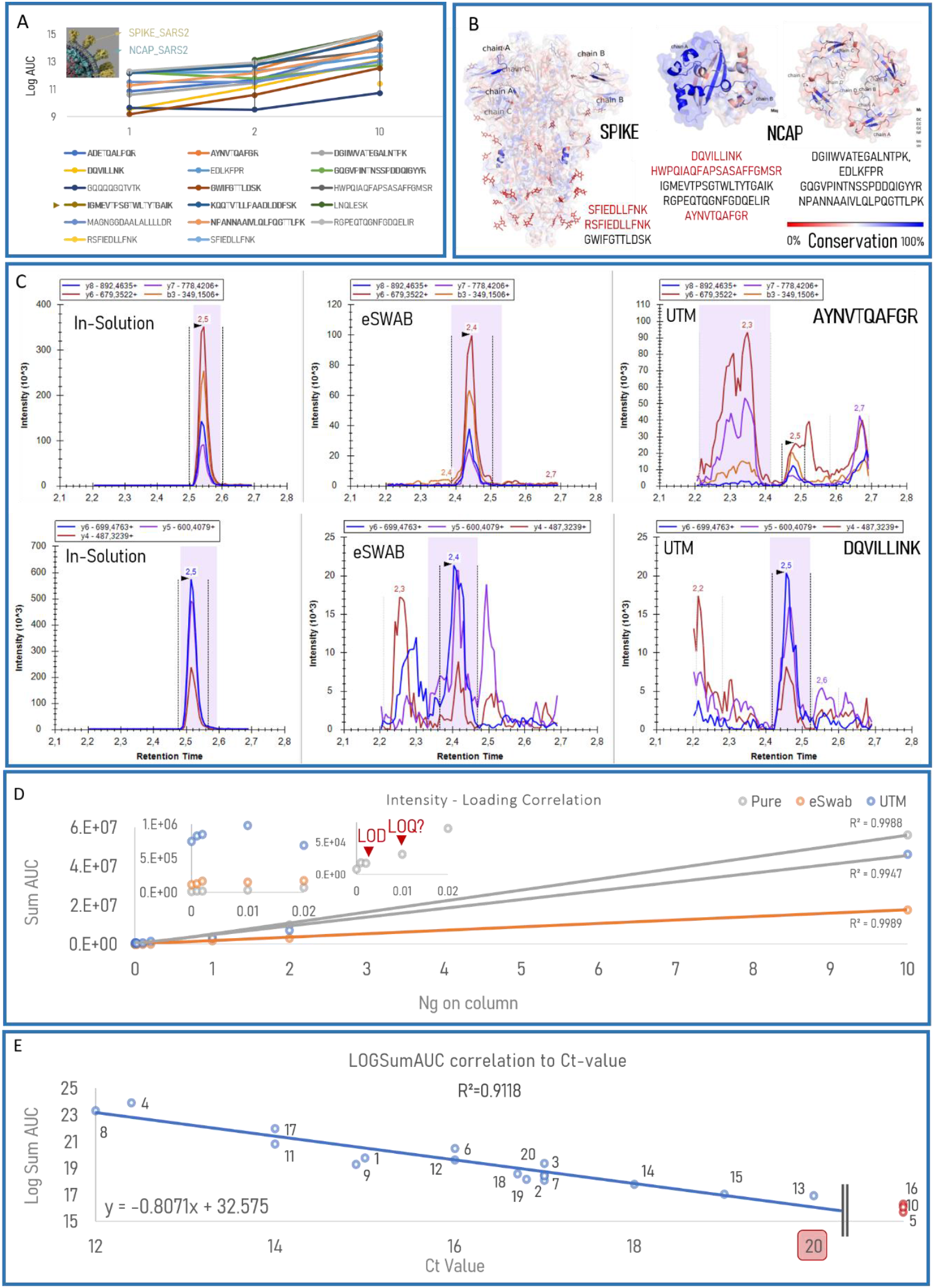
Discovery phase (Blue). (A) Seventeen SARS-CoV-2 responsive peptide biomarkers. Based on public data (16,21), we selected two target proteins (NCAP_SARS2 and SPIKE_SARS2) and obtained recombinant equivalents (Sino Biological). Using dilution series in a background of 250 µl UTM from nasopharyngeal swabs of healthy volunteers, we used our recently published workflow combining narrow window DIA with predicted intensities and retention time to detect a total of seventeen responsive peptides (30). Only intensities of peptidoforms that passed the mProphet threshold for 1% FDR are depicted. The arrowhead indicates a substituted amino acid in the sequence in the recombinant protein. Bold sequences were also reported in a recent review from Grossegesse M. et al. (17). **(B) Peptide mapping and evolutionary conservation**. 12 out of 17 peptides were mapped onto SPIKE_SARS2, NCAP_SARS2 C-terminal dimerization domain and NCAP_SARS2 RNA binding domain (from left to right). Mapped peptides are highlighted in structures and the conservation scores were colored red (0% conserved) to blue (100% conserved) scale. Black font indicates sequences unique for SARS-CoV-2. **(C) Matrix interference and peptide selection**. Irrespective of their evolutionary conservation, all candidate peptides were retained to allow robust assay development compatible with any matrix, experimental condition or LC-MS instrument used. Depending on the sample conservation buffer, intensities and interferences vary greatly, as illustrated by the transition signals from AYNVTQAFGR and DQVILLINK at 2.5 minutes without (in-solution) and with an eSwab and UTM background. Arrowheads indicate correct peak. Signal within dotted lines, i.e. peak boundaries, is summed to calculate the LogSumAUC. **(D) Intensity – loading correlation in three different backgrounds**. The recombinant proteins were measured in different loading amounts either without matrix (in-solution) or in eSwab (saline matrix) or UTM (protein matrix) (5 µl medium-equivalent oc). Left inset illustrates the impact of the background on low intensity signal. The right inset highlights the instrumental Limit of detection (LOD) and potential limit of quantification (LOQ) of these viral proteins in-solution when all transition intensities are summed. **(E) Correlation between MS protein signal and RT-PCR RNA detection**. Starting from the Skyline document, an MRM was developed for UTM samples on a Xevo TQ-S instrument (Waters), by MRM transition selection. The final method comprised ten peptides with a total of 30 transitions. We applied the MRM assay to the twenty patient samples in UTM obtained from a University Hospital (Leuven). An equivalent of 5 µl (out of 3 ml) of medium of each sample was loaded oc. The detection results from the Skyline report were used to logarithmically transform the summed AUC (LogSumAUC) of all the peptides. When plotted against the Ct value measured by RT-PCR, a strong correlation is found (formula allows conversion of Ct into expected signal), which suggests that this assay can have great potential in the clinic. Negative patients (red) are depicted by their LogSumAUC only.

#### Translation into a targeted MRM assay with instant clinical applicability

As a template for all downstream optimizations, a Skyline project was created on the DIA data, retaining all seventeen peptide biomarkers with 145 transitions (**Supplementary data 4**). Notably, two peptides with a missed cleavage were retained because they derive from a KK, RR, KR or RK amino acid sequence motif (35). To avoid false positive results, however it is important to verify that biomarker peptides are taxonomically uniquely assigned to SARS-Cov-2 (**Supplementary Data 5**), and to understand the evolutionary conservation of these peptides (**Figure 2B**). Our taxonomic analysis using Unipept (**Supplementary Data 5**) illustrates that some of the detected peptides are also expressed in other *Coronaviridae* or even different organisms. While others have suggested to discard all non-unique sequences during target selection (15,29), we maintained these for downstream optimizations. First, a combination of peptides allows higher specificity (17) and to account for intrinsic incompleteness of databases. Second, the prevalence of other organisms in nasopharyngeal swabs is not known, and these peptides may therefore never occur apart from SARS-CoV-2 infection. Most importantly, however, compared to RT-PCR, implementation of viral detection MRM assays in the clinic is much more limited by the variability in sampling background (e.g. blood and mucus), conservation medium and instrument specifications. Therefore, the intensities, retention times (t_R_) and interferences of transitions between different samples can vary greatly when the matrix is not removed. To illustrate this, we depict the signal of AYNVTQAFGR and DQVILLINK, both at a t_R_ of 2.5 minutes, as these were measured without matrix (in-solution), in UTM (dominant protein background) and in eSwab medium (saline background) with 1 ng on-column (oc) (**Figure 2C**). In conclusion, we recommend targeting all seventeen peptides as a starting point in every unique setup where the assay will be implemented. The rollout of an MRM assay is then reduced to omitting those MRM transitions with the poorest performance for that site.

Using this Skyline document, the first MRM assay was optimized on a Xevo TQ-S (Waters Corporation, Wilmslow, United Kingdom). It contained ten peptides, measuring 30 transitions (**Detailed Methods Section and Supplementary Data 6 and 7). Figure 2D** shows that summing the intensities of these 30 transitions (irrespective of individual peak shapes or signal-to-noise) provides a robust metric to quantify dilution series of SARS-CoV-2 proteins with and without matrix. However, the preservation medium has a considerable impact, as shown by background inflation of low signals (in-solution < eSwab < UTM, Figure 2D left inset) and by matrix suppression of more abundant signals (in-solution > UTM > eSwab). Notably, SPIKE could still be detected down to 70 amol (10 pg) oc and NCAP down to 40 amol (2 pg) oc in solution. This is referred to as the instrumental Limit of Detection (LOD) (**Figure 2D right Inset and Supplementary data 6**). In fact, LogSumAUC still allows to distinguish 0.001 and 0.002 from 0ng oc, while no clear peaks could be annotated in the Skyline project. This implies that summing intensities at specific retention times could be a more robust metric than trying to call single peptide peaks. At 10 pg oc, the summed intensity of all transitions was higher than that of 2 pg, making this the potential instrumental limit of quantification (LOQ). Assuming 300 NCAP molecules in a viral particle, as few as 80.000 particles can thus theoretically be detected without matrix, in line with previous reports (9,10,36).

Finally, this optimized MRM assay was applied on the twenty UTM patient samples obtained from University Hospital Leuven, which had been previously diagnosed using RT-PCR. The RT-PCR result is measured by the number of RNA duplication cycles required to detect viral RNA, designated the Ct value. Therefore, peak boundary corrected transition intensities were summed and Log2-transformed (LogSumAUC) and correlated with the RT-PCR Ct values. From these 5 µl UTM oc LC-MS experiments, a strong inverse linear correlation (R^2^ = 0.9118 with the three negative samples excluded) was found between the LogSumAUC from Skyline and the Ct values measured in the clinic (**Figure 2E, Supplementary data 7**). This strong correlation was already suspected (17) and makes an overwhelmingly strong case for the diagnostic potential of the protein-based MRM assay.

### The Cov-MS consortium (Red): Increasing MRM assay sensitivity and robustness

Based on the observed linear correlation between the measured peptide intensities and the RT-PCR Ct values (**Figure 2E**), the LOD of the assay, i.e. its sensitivity, was redefined in the context of Ct values measured in the clinic. This comparison allows improvements to the assay to be expressed in terms of number of Ct cycles gained, with one Ct cycle corresponding to a doubling in sensitivity. Indeed, for a larger batch of patient samples (n = 86; 5µl medium equivalent oc) from another hospital (AZ Delta) and that included higher Ct values and different media (**Supplementary data 8**), the decrease in LogSumAUC stalled around Ct 20 and there was no difference in measured signal-to-noise (S/N) at higher Ct values. Therefore, the next challenge was redefined as increasing the S/N in order to gain as many Ct values as possible, i.e. to gain detection doublings. In essence, this implied straightening of the curve (**Figure 3A**).

**Figure 3.**
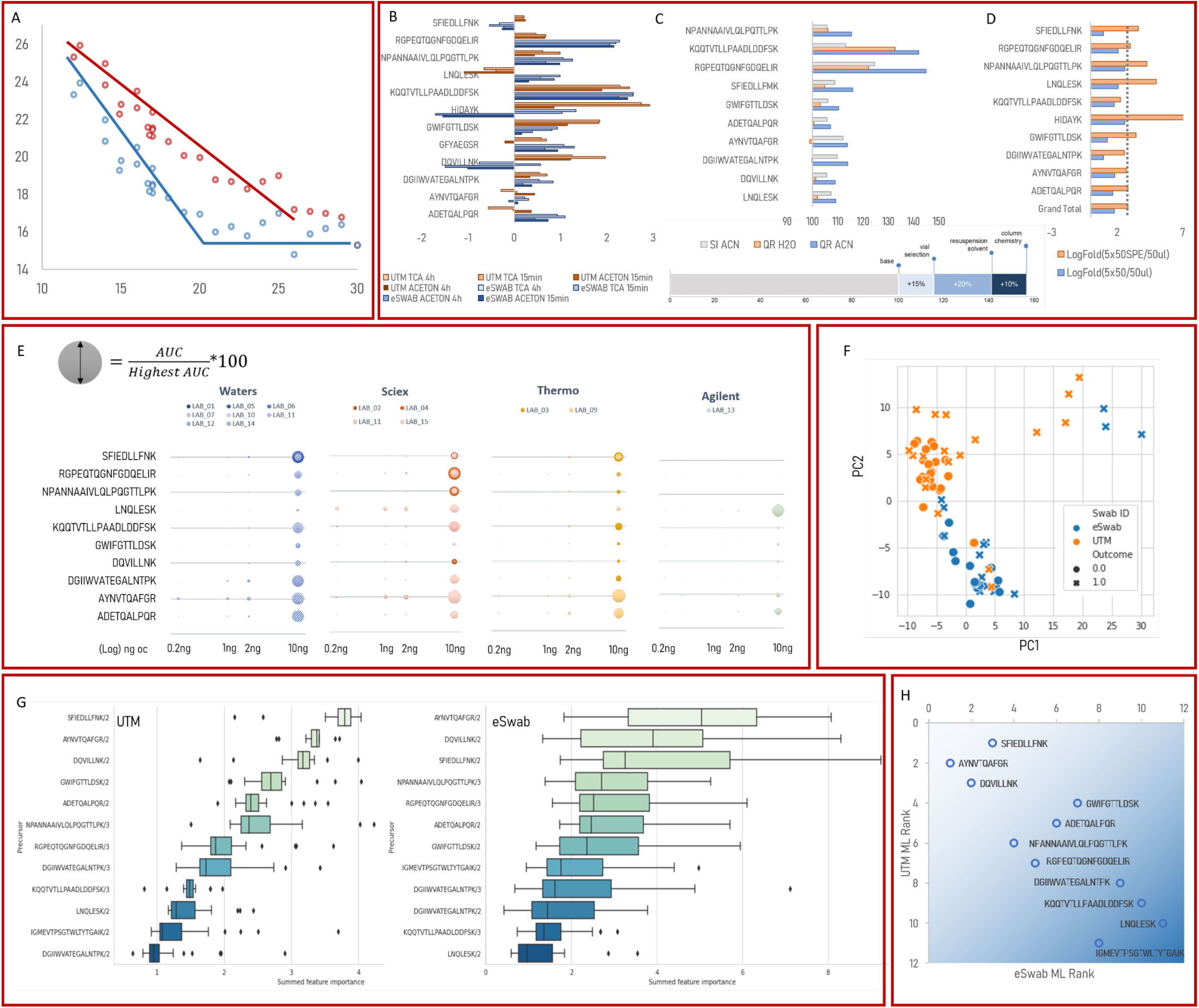
The Cov-MS consortium report (Red Phase). **(A) The aim of the consortium** was to increase the signal to noise from the discovery phase MRM assay (blue, **Supplementary Data 7 and 8**) to “straighten the curve” (red) (cartoon representation). **(B) The sample preparation optimization**. Different storage media, TCA precipitation and a 15 minutes digest (n = 5) were compared. The increased LogSumAUC is expressed as log fold changes. **(C) Comparing different sample preparation consumables**. In the standard workflow peptides were dissolved in water and transferred to spring insert (SI) sample vials. Using Quan Recovery (QR) vials and adding 5% acetonitrile (ACN) to the solvent increased the signal for most peptides in solvent, but not in UTM (**Supplementary data 10**). The bottom bar displays the estimated overall gain attributed to consumables. **(D) Increasing the sample load for eSwab samples**. Because eSwabs use saline-based preservation solution, increasing the amount of sample oc can be beneficial. Therefore, we assessed the impact of increasing the loading fivefold, either with or without SPE. An overall average LogSumAUC fold change of 2.3 (dotted line), i.e. 5 times increase in signal, can be attained if the sample is split in five and SPE is applied (LogFold 5×50SPE). **(E) Data acquisition within the Cov-MS Consortium**. 15 labs optimized acquisition on their instrumental platforms using the SOP, the Cov-MS kit and the Skyline project provided. The bubble plot represents the relative abundance of each peptide in the dilution series compared to the highest abundant peptide for that lab. Strikingly, each lab detected a different (collection of) peptide(s) as the best targets. Several labs could detect signal down to 0.2 ng oc in UTM background. LAB_14 is depicted with shading because this instrument was fitted with a UniSpray Source. **(F) Optimization of the data analysis**. We developed a more data-driven scoring function as an alternative to simple summation of all signal (LogSumAUC). Therefore, a model was trained using all features exported from the Skyline document of 70 patient samples from the AZ Delta sample batch. The principal component analysis (PCA) scoring the distribution of all features, illustrates that the conservation medium is one of the most prominent variables in the dataset. **(G) Peptide feature weighing**. The feature weights that are given to each of the MRM transitions by the ML algorithm are representative of their diagnostic value. This in turn can be used to calculate the contribution of each peptide to diagnostic outcome. **(H) A final peptide ranking in two different media**. From the upper left to the lower right, peptides are depicted by their ML rank in both media. Three peptides could be universally applicable (on a Xevo instrument), irrespective of the medium.

On April 21, 2020 we presented our discovery phase and preliminary results online to over 80 interested parties to establish a consortium (**Cov-MS**) to broaden the scope of the work, transfer the methods to other (vendor) instruments, and bring in the expertise of other functions and scientists (37). This allowed parallelization of the sample preparation, data acquisition and data analysis optimization (**Figure 1 a-c**).

#### Sample Preparation optimizations: 2-3 Ct values

Efforts focused around three optimizations: applying a different precipitation strategy with acid (TCA and HCl), reducing digestion time to fifteen minutes, and using different conservation media. While many of these changes increased overall signal (**Figure 3B, Supplementary data 9**), the best protocol is typically dependent on the MRM assay under development as many changes are peptide-specific and might be attenuated by increased ion suppression or noise depending on the medium, sample related specifics, and experimental conditions. Nevertheless, the reduction of the digestion protocol to only fifteen minutes now enables a complete sample preparation workflow of less than 30 minutes.

In parallel, the use of different LC-columns, sample vials and resuspension protocols were also investigated, alongside peptide stability (**Figure 3C, Supplementary data 10**). Beneficial effects were incremental, on the order of 10-20% gains in LogSumAUC for (i) using 5% acetonitrile for redissolving the peptides, (ii) using dedicated sample containers (QuanRecovery Vials, Waters Corporation, Milford, MA), and (iii) reversed phase separation columns to minimize non-specific binding (ACQUITY PREMIER Peptide BEH C18 Column, 300Å, 1.7 µm, 2.1 x 50 mm, Waters Corporation, Milford, MA). Notably, the gain is lower in UTM background, most probably due to carrier protein effects in this medium.

Crucially, the biggest limitation in terms of sensitivity is the amount of sample that can be loaded oc. More specifically, the results from the discovery phase were all attained on 5µl medium equivalent oc. Unfortunately, increasing sample load is not straightforward for MRM assays. In fact, UTM was designed to enable post-sampling culturing of pathogens and therefore contains large amounts of collagen and bovine plasma proteins. This protein background negatively impacts the assay in three different ways: (i) it can cause severe MRM transition interferences that can increase false-positive rates and hamper automated peak calling (**Figure 2C**); (ii) certain peptides suffer from ion suppression (**Figure 2C**), and (iii) most importantly, we experienced carry-over and instrument contamination due to the presence of collagen (**Supplementary Data 10**) (17). Combined, these factors undermine LC-MS assay robustness. eSwabs, on the other hand, are preserved in 1 ml (as opposed to 3 ml) of saline buffer without protein background. These could allow loading 5 to 50-fold more sample oc without compromising instrument stability. This means that e.g. 250 µl (instead of 5 µl) of the initial 1 ml of eSwab can theoretically be used for a single injection, which is comparable to RT-PCR sample consumption and will easily accommodate subsequent re-analysis of the same sample. The precipitation step in turn allows to concentrate the sample without drying. However, we noticed that handling the 250 µl acetone precipitate directly in 2 ml vials was causing unexpected sample losses and that loading 50 µl of sample in a single LC-MS injection interfered with retention of early eluting peptides. Therefore, we compared the following three protocols on 50 µl starting volume and with 5µl injection: (i) 50 µl of eSwab medium precipitated and resuspended in 50 µl digestion buffer (5 µl medium equivalent oc), (ii) five times 50 µl precipitated, each digested in 10 µl and merged back into 50 µl digestion buffer (25 µl medium equivalent oc), and (iii) five times 50 µl precipitated, each digested in 50 µl buffer and concentrated together onto a single solid phase extraction (SPE) column and eluted in 50 µl (25 µl medium equivalent oc). The fold changes of each peptide are shown in **Figure 3D (Supplementary Data 11**). The anticipated five-fold increase in signal (dotted line), i.e. minimum two Ct values, can be attained if five times 50 µl digest solution is concentrated onto a single SPE column. Automation will allow this finding to be consolidated and extended, yet we caution here for SPE overloading. Given the lack of in-house automation, we continued with the original protocol of 50 µl and 5µl injection without applying SPE.

#### Data acquisition optimization: 1-2 Ct values

After the online presentation of the assay on April 21st, 2020, many of the attendees joined the Microsoft Teams Cov-MS Team containing three channels, one per participating vendor at that time (Waters Corporation, Sciex, and Thermo Fisher Scientific). At the time of submission, the group counted over 100 participants. The Cov-MS labs were encouraged to discuss method optimization with the vendors’ international expert support teams.

To streamline the integration of all these parallel optimizations, consortium members were provided a detailed Standard Operating Procedure (SOP) for method optimization, the Skyline template containing the seventeen peptides (145 transitions) and a Cov-MS Kit (**Supplementary data 12**). These kits contained digests of pure recombinant NCAP and SPIKE protein as well as a dilution series in a UTM background from healthy donors, in triplicate. The kits allow members to standardize the optimization protocol, to assess performance under different circumstances, and to centralize all data to distill best practices per vendor. **Figure 3E** summarizes overall peptide detectability for fifteen participating labs. Individual instrumental platform data is provided in **Supplementary data 13 and 14** in an easily handled pivot table format. Several partners compiled detailed reports, and Sciex and Waters Corporation published several application notes based on their data (38,39) (**Supplementary data 15**).

Importantly, only the latest generation tandem quadrupole instruments can reach adequate sensitivity. Additionally, the compiled results confirm the initial hypothesis that peptide selection is lab-specific. This was recently also described in a review on currently available MS-based detection assays (17). However, because the data in this study were acquired on the same samples and analyzed using the same software, it is possible to draw direct conclusions on the acquisition itself. These preliminary results imply that different vendors could benefit from different sets of target peptides, even if the same transitions were targeted (**Figure 3E**). One striking example is the RGEPQTQGNFGDQELIR, which is especially prominent on Sciex instruments, possibly because the corresponding ion sources are better suited to higher charged peptides. The impact of the source is best illustrated by LAB_14 that used a UniSpray source (Waters Corporation) instead of an ESI source, which boosts the ionization of the ADETQALPQR peptide compared to all other labs. The latter is especially interesting in light of a previous report that argues for this target peptide (17,40). While this remains to be fully confirmed by the growing Cov-MS consortium and beyond, including ionization efficiencies in MALDI sources, the drafting of vendor-specific best practices will likely be important for streamlined lab-specific assay rollout.

#### Data Analysis optimization: 1-2 Ct values

**Figure 2E** depicts the quantitative MRM results as the LogSumAUC of all ions within the peak boundaries of the transitions in that particular assay. The initial selection of these transitions was based on an expert’s choice from the UTM dilution series in the Cov-MS kit. However, not all transitions contribute to diagnosis equally and leaving out those transitions that are subject to interference in a specific matrix, have low signal, or are simply redundant to accurately detect a peptide, allows either retention of only the best transitions in the final LogSumAUC, or increased instrumental dwell times to boost sensitivity. However, because of the high variability between samples, this optimization can only be attained by a data-driven approach.

To construct such a data-driven scoring function to distinguish between positive from negative COVID-19 patients from the MRM data, a machine learning (ML) model was trained and evaluated (**Detailed Methods section**). The training and evaluation data consisted of a Skyline document export of 70 LC-MS experiments on a Xevo TQ-S (Waters Corporation) accompanied by relevant metadata (healthy or diseased and medium used), Ct as determined by RT-PCR at the time of initial diagnosis (**Supplementary Data 8**). The full script and the weights of the individual transitions is available in **Supplementary Data 16 and on github**.

The principal component analysis of the feature scores exported from Skyline clearly illustrates that the medium is the most prominent contributor to the second principal component and thus to the choice of transitions and the attainable diagnostic value (**Figure 3F**). Therefore, in a second round, we trained the two patient populations separately (eSwab n = 29 and UTM n = 41). **Figure 3G** depicts the diagnostic weight of each peptide in the respective media. Fortunately, a common set of three peptides are well suited for both UTM and eSwab, covering both SPIKE and NCAP structural proteins, making this a very attractive target selection (**Figure 3H**).

### Towards a clinical MRM Assay (Green)

From the start, our joint efforts have focused on fast and efficient clinical applicability. Therefore, we retain the matrix, reduce sample preparation to below 30 minutes, provide kits, SOPs and software methods, and only use tandem quadrupole instruments in configurations that are readily available in the clinic. Finally, we describe the first steps in standardization and validation of the assay for clinical implementation and present our Cov-MS Digital Incubator to facilitate communication and exchange on further improvements.

#### The Cov-MS internal standard assesses sampling efficiency, sample preparation, data acquisition and data analysis automation

In clinical MS-based assays, stable-isotope-labeled (SIL) internal standards are indispensable to meet measurement accuracy and to satisfy regulatory requirements. For this assay, we designed the Cov-MS QConCAT construct (PolyQuant) (41) (**Figure 4A**). It not only allows to report absolute viral load and interlaboratory comparison, it also enables validation of the entire workflow: (i) swab sampling procedure efficiency in the clinic, (ii) sample preparation efficiency and (iii) instrument stability, all whilst (iv) greatly enhancing automation potential of the data analysis. Briefly, PolyQuant (Bad Abbach, Germany) joined the Cov-MS consortium and initiated the expression of a QConCAT protein construct in *E*.*coli* that contains the seventeen peptide biomarkers. These are heavy labeled through the incorporation of ^15^N and separated by their native amino acid context to assess the efficiency of the digest. Additionally, three RePLical retention time standard peptides were incorporated to help assess LC stability. Finally, four peptides, one from each of the human core histone proteins, were added to the labelled construct to serve as host protein markers for swab sampling quality. We reasoned that nuclear proteins will only be present if the swab caused (superficial) tissue damage to release intracellular viral particles from cells. Therefore, when histones are low or absent, this could point towards poor swab sampling efficiency and thus a potential false-negative result. Indeed, all twenty patients from **Figure 2C** contained at least one of these four histone peptides in the DDA runs (**Supplementary data 3C and 17**). Therefore, the MS-Cov QconCAT construct will help in end-to-end assay troubleshooting, and enables the direct comparison of results from all over the world.

**Figure 4.**
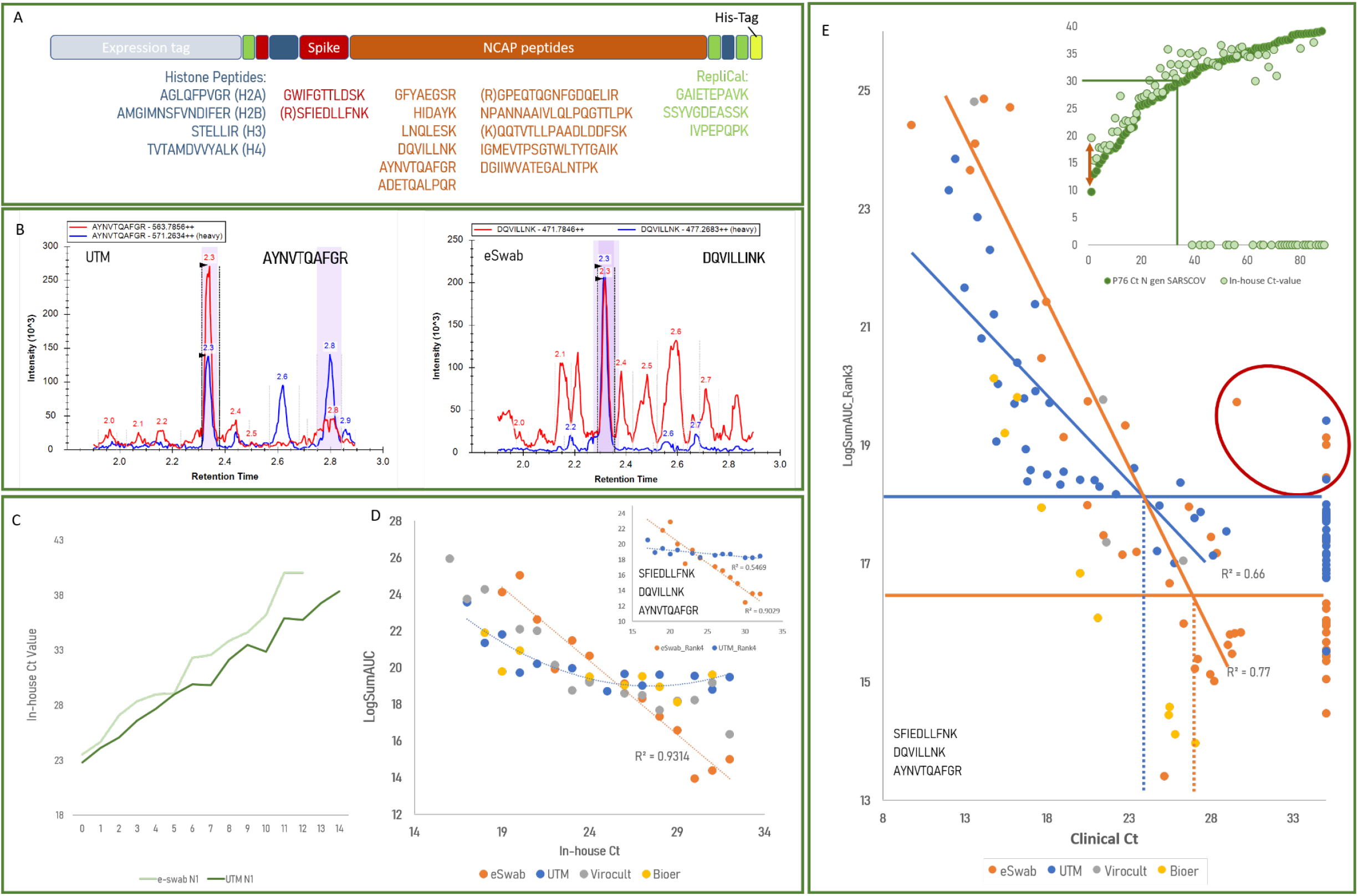
Towards translating the Cov-MS assay into the clinic. **(Green Phase) (A) The Cov-MS QConCAT internal standard**. An isotope labeled (‘heavy’) construct was expressed in *E. coli* to troubleshoot and standardize assay development. It contains the seventeen Stable Isotope Labelled (SIL) SARS-CoV-2 peptides for absolute quantification (red and orange), three RePLliCal peptides to assess LC system stability (green), and four host peptides derived from histones to assess the efficiency of the swab sampling procedure (dark blue). **(B) Facilitating peak detection using the Cov-MS internal standard**. The two peptides from Figure 2C are depicted in red in their most interfering matrix. The blue trace depicts the heavy signal from the Cov-MS SIL (2ng and 2.5ng oc respectively), showing how peak detection now becomes automatable. **(C) in-house Ct-values for a dilution series of viral particles from the National Reference Center (NRC) in Belgium**. We generated dilution series in two different backgrounds and determined the Ct values using our In-house RT-PCR assay. eSwab consistently gave higher Ct values compared to UTM in our hands, but the detection limits attained would allow for accreditation of our RT-PCR workflow. **(D) Patient Dilution series as an Alternative Assessment Procedure (AAP)**. The viral particle used in the dilution series presented in (C) did not yield any reliable signal in the MRM assay. Therefore, four patients with comparable in-house Ct values in four different backgrounds were selected and diluted in their respective background of negative patients. The X-axis depicts the theoretical Ct values in such dilution series according to (C) and the y-axis represents the LogSumAUC of the 13 peptides retained in this MRM assay **(Supplementary Data 19)**. The Cov-MS QConCAT was not yet included in this experiment. Most notably, eSwab retains a linear correlation with intensity below a LogSumAUC of 18 which is not noticeable in the other media. The inset shows the results when only the best three peptides from the ML (**Figure 3G**) are used for eSwab and UTM. **(E) A large patient cohort**. Inset: 89 clinically positive patients (dark green) were first selected for in-house RT-PCR (light green) and both Ct values were compared. Negative in-house results are depicted on the X-axis. The arrow (inset) highlights the discrepancy between the eSwab clinical and in-house Ct values for the Patient used for the dilution series in (D). All patients with a clinical Ct below 30 were positive in both assays and were retained for MRM assay validation (n = 82). Using the best three peptides from the ML (**Figure 3G**), the LogSumAUC was plotted against the clinical Ct value. Negative patients are depicted to the right to sample the noise. The outliers are circled in red. The horizontal line shows the LogSumAUC_Rank3 below which signal cannot be confidently distinguished from noise in UTM (blue) and eSwab (orange). For both media the cross-section with the linear regression line is projected downwards to estimate the theoretical sensitivity in terms of Ct value.

A Skyline document comprising both ‘heavy’ (from the QcontCAT protein) and ‘light’ (from the native viral and human proteins) transitions was compiled and three oc loadings were compared for both UTM and eSwab (**Supplementary Data 17). Figure 4B** shows the compiled signal of the light and heavy form of the two peptides from **Figure 2C** in their most interfering matrices. It is clear that the heavy signal (detected here by one transition) greatly facilitates correct peak detection. The discussion on peak detection automation in Skyline can be followed in the Skyline support discussion thread (42). Notably, 1.25 ng was loaded in the final patient batch to avoid native peptide ion suppression, but this turned out not to be enough, especially in UTM background. Additionally, it might prove beneficial in the future to target more transitions of the heavy and light peptides to facilitate peak detection and retain the *dotp* value as an additional feature in the Skyline report. Finally, we noted that nearly all peptides (from the QConCAT) display a forward retention time shift in UTM background compared to eSwab (**Supplementary Data 17**).

#### RT-PCR Accreditation Standard and Ct values are not easily transposable to an MRM assay

For external quality assessment (EQA), we requested positive control material from the National Reference Centre (NRC) in Belgium. Developed for RT-PCR, this purified heat-inactivated virus is used to create a dilution series that allows labs to determine their LOD. These are series of ½ dilutions, each thus corresponding to one Ct cycle. The starting solution (dilution 0) was said to be the equivalent of a Ct17 in the hands of the NRC (**Supplementary Data 18**). By simply applying the formula of **Figure 1E**, a linear correlation with their patient Ct values would allow us to easily detect the first few dilutions, i.e. by a predicted LogSumAUC of 19.3. However, because different RT-PCR assays can vary up to orders of magnitude in the number of copies that can be detected (3), we first developed an in-house RT-PCR assay using plasmids as targets. Applying the manufacturer’s instructions, we were able to detect down to ten copies of the N-gene plasmids at a Ct of 38, which corresponds to 2.5 copies at Ct40, the theoretical upper limit for RT-PCR diagnosis (**Supplementary Data 18**).

We then measured this NRC dilution series in negative patient background with our in-house RT-PCR assay and could detect the N1 gene after ten (Ct 36) and fourteen (Ct 38) ½ dilutions in eSwab and UTM, respectively, which would allow for accreditation (**Figure 4C and Supplementary data 18**). However, this still was 5-6 Ct values higher than in the hands of the NRC. Additionally, several attempts to detect peptide signal in these standards by means of LC-MS failed. This is most surprising, given the fact that the patients in **Figure 2D** were analyzed with 5 µl medium equivalent oc and that we now loaded up to 100µl medium equivalent for these standards using an optimized acquisition and sample preparation strategy. We attribute this outcome to (a combination of) any of the following reasons: (i) sample degradation (both mRNA and protein), (ii) specific sample preparation steps being incompatible for the specific purpose of detecting proteins in these viral particle standards, (iii) the heat inactivation step or (iv) the potential role played by viral-like particles (VLP) (18,19), which were not present in these ultracentrifuge purified standards. In other words, in patient samples, a higher signal for proteins could be present than would be theoretically anticipated from the RT-PCR positive control set. This implies that proficiency assessment schemes for this assay will only be possible through an Alternative Assessment Procedure (AAP), as is often the case when different biomolecules are targeted in clinical assays (See section 9.1.1 from CLSI 62-A).

We therefore propose patient dilution series as an AAP. More specifically, a dilution series of positive patient samples in the respective sample buffer from negative patient samples allows assessing the sensitivity of the optimized MRM assay in terms of Ct value. We made four such dilution series in four different matrices, now also including Virocult and Bioer. We selected four patients with a comparable in-house Ct value and based on **Figure 4C** we assume that every ½ dilution increases the Ct value by one. **Figure 4D** shows the LogSumAUC of all peptides, because no ML has been done on Bioer and Virocult at this point. Most notably, the eSwab signal retains its linear correlation to Ct value down to a much lower signal, which is indicative of a lower background signal, i.e. lower noise and interference compared to the other media (**Supplementary data 19**). In fact, when only the rank three peptides for UTM and eSwab are used to present the results, the linearity in UTM persists as well, albeit at a much shallower slope, implying that the ML possibly has selected for peptides with fewer interferences (**Figure 4D inset**).

Note that our in-house RT-PCR consistently has a higher Ct value in eSwab compared to the UTM (**Figure 4C**) and that the patient that was used for the eSwab turned out to have a large discrepancy between the in-house and clinical Ct, even after a second run (arrow in **Figure 4E**). Additionally, we have been presenting the strong correlation between Ct and LogSumAUC as a measure to assess the performance of an MRM, yet such correlation does not need to be linear in patient batches. In fact, a large discrepancy in MRM signal between two patients with identical Ct values could have relevant biological grounds, e.g. be an indication of an infectious or “superspreader” phenotype, correlate with the symptomatology and/or with (anticipated) severity of disease. We therefore argue against setting a specific Ct value as a threshold for MRM assay sensitivity and emphasize that the focus of interpretation of these results should be on the low noise level in eSwabs, rather than the actual Ct values.

In conclusion, a solid validation of this AAP is required before it can be universally adopted. In fact, measuring increasingly large patient cohorts, ideally along with documentation of (absence or presence of) symptoms could prove more useful to validate the performance of an MRM assay in a clinical setting.

#### A large patient cohort for assessing potential assay performance

As a final validation of the current performance of our Cov-MS assay, we analyzed 325 patient samples with the optimized conditions from the Cov-MS collaboration on a Xevo TQ-XS instrument (Waters Corporation) (**Supplementary Data 20**). To be able to compare the four different media in this batch, we prepared only 50 µl of each sample, followed by precipitation and resuspension in 25µl digest buffer and 10 µl injections, i.e. 20 µl medium-equivalents oc. In parallel, we diagnosed 89 positive patients in the batch with our in-house RT-PCR. Remarkably, at lower Ct values we consistently measured a higher than expected Ct value. At higher Ct values (> 30) many of the patients did not even yield any signal for the N gene in-house (**Figure 4E inset**). This can be explained in part by the fact that the clinical RT-PCR was done on more sample. More specifically, the starting (190 vs 140 µl), elution (40µl vs 60µl) and final input volume (8 vs 5 µl) together make that our in-house assay only uses 30% of the volume in the PCR reaction, i.e. ∼10 µl, compared to the clinical assay, corresponding to nearly 2 Ct values. Additionally, these samples had been preserved in the freezer at −20°C for over four months. Finally, low positive patients can be expected to be more prone to drop-out in the higher Ct range, i.e. after a larger number of amplification cycles, wherein error too can accumulate. Therefore, we only used patients with clinical Ct values below 30 as positive and with negative clinical diagnosis as negative. Note that this high degree of variability between RT-PCR assays again impairs an easy definition for specificity and sensitivity of the MRM assay based on a Ct threshold.

**Figure 4E** shows the final outcome of the MRM assay using the rank 3 peptides from **Figure 3G** on the remaining 82 positive patients, i.e. 37 positive patients in UTM, 30 patients in eSwab, 11 patients in Bioer and 4 patients in Virocult. 56 negative patients in eSwab (22) and UTM (34) were subjected to the same manual peak boundary curation to define the background signal. Notably, the Cov-MS QConCAT construct was not detectable in all samples because the standard turned out to be underloaded at 1.25ng oc and was only used to facilitate manual setting of peak boundaries. Thus, 138 out of the 325 patients were used in the final data analysis of the large patient cohort. Raw data of all runs is available on the Skyline Panorama server. As an alternative to setting a hard Ct threshold, the minimum background threshold is highlighted for UTM and eSwab and the intersection with the linear regression of the positive patients is projected onto the Ct axis. This is the Ct above which the background, i.e. matrix, becomes too high to distinguish signal from noise and we currently consider this to be the theoretical maximum sensitivity using the current workflow. All patient samples were analyzed in a randomized sample list (**Supplementary Data 20**) and at least some of the background outliers of eSwab could be attributed to the fact that they were run closely following a UTM sample and were subject to carry-over. In conclusion, **background, i**.**e. matrix, and not instrumental limitations, is currently the limiting factor for increasing the sensitivity**. In fact, the red line of the “straighten the curve” cartoon in **Figure 3A** should be imagined continuing down below the blue curve…

We conclude that saline buffers are the only matrices that allow easy adaptation of the assay and to diagnose patients with Ct values above twenty without matrix removal. In fact, we strongly argue against the use of the protein-laden UTM for MS-based SARS-CoV-2 diagnosis given the reasons mentioned before. **In brief, the maximum attainable single-shot MRM would thus start with direct ice-cold aceton precipitation (7 volumes) on 175 µl of saline medium in 1**.**5 ml Eppendorf vials, resuspending this in 35 µl 50 mM TEABC 0**.**5 µg trypsin/LysC and 5% acetonitrile with 10ng Cov-MS QConCAT standard, digesting for 15 minutes at 37°C, followed by addition of 3**.**5µl of 10% formic acid (end concentration 1%) and - possibly following SPE - injecting 10 µl oc**. This is **45**.**5 µl medium-equivalent oc**, or 2.25 times more than the patient batch analyzed here. **While it is currently not proven that increasing the loading increases sensitivity - as the matrix background raises accordingly** - we are confident that Cov-MS has strong potential of becoming a valuable addition to the diagnostic SARS-CoV-2 toolbox in the near future.

#### The Cov-MS Digital Incubator: clinical roll out and facilitating improvements

Taken together, the high degree of variation in matrices and instrumentation was intercepted in this study by collaboration in the Cov-MS consortium. Uniquely, both academic and industrial partners have shown inspiring openness in a joint effort to quickly extend testing capacity and help alleviate pressure on current testing facilities. However, the current assay cannot be directly implemented into the clinic. Rather, it is an academic end point and initiatives like COVID-MSC can now take over and move into the next phase (23). In fact, in both the UK and the Netherlands institutionally driven efforts have been initiated. In order to facilitate this, we therefore propose to persist the collaborative nature of this effort and we invite all interested parties with MS capabilities to join the Cov-MS Digital Incubator Microsoft Teams Environment, so they can access all information gathered during this initial phase of development, and can parallelize further efforts to improve applicability, robustness and sensitivity of the assay. Access can be requested by sending name and affiliation to.

### Future perspectives

We provided a detailed description of a community effort to build a robust and sensitive orthogonal assay to diagnose SARS-CoV-2 positivity. To be able to track and align with the millions of RT-PCR tests that are performed daily, a collective effort by the scientific community is paramount, not in the least because MRM assays have rarely been used before to detect viruses (17). Therefore, we invite other labs to build on this platform and to introduce further optimizations across all parts of the workflow to help translate this assay into mainstream clinical laboratories and even to potentially start adding other pathogens to the assay. Here, we list some of the most pertinent issues and propose some potential solutions.

#### Increasing Sensitivity

**Figure 5A** displays the calculations that allow to discuss future improvements in terms of absolute numbers. Briefly, a perfect RT-PCR assay (as was done in-house on plasmids, **Supplementary Data 18**) detects 10 genomes in 10µl at a Ct of 38, i.e. 2.5 genomes at Ct 40. With every virion carrying 300-350 NCAP molecules (36), the absolute amount of NCAP (in attomole) in 10µl is easily calculated. Therefore, the question becomes what can still be detected by current instruments. We distinguish three different thresholds (**Figure 5B**): (i) the signal attained in the current assay, i.e. in eSwab background with a QConCAT for efficient peak picking of low individual signals, (ii) pure peptides, i.e. how individual signals could look like following enrichment and (iii) what might be attainable by ML-approaches or if signal is simply summed and no single peaks are required (**Figure 1D and Supplementary Data 21**). The latter strategy needs to be further validated to assure its applicability. Still, **Figure 5C** depicts increased Ct values to illustrate what could happen if (a) target peptides are first enriched and up to 320µl is used in a single shot (32 times more, i.e. 5 Ct values) and/or (b) patient Ct values are higher in clinical labs compared to the perfect plasmid situation and/or (c) the potential presence of viral-like particles boosts the signal on an MRM assay. Based on these calculations, we predict that a clinical Ct value of 30 will most probably be the maximum attainable for current mass spectrometry.

**Figure 5.**
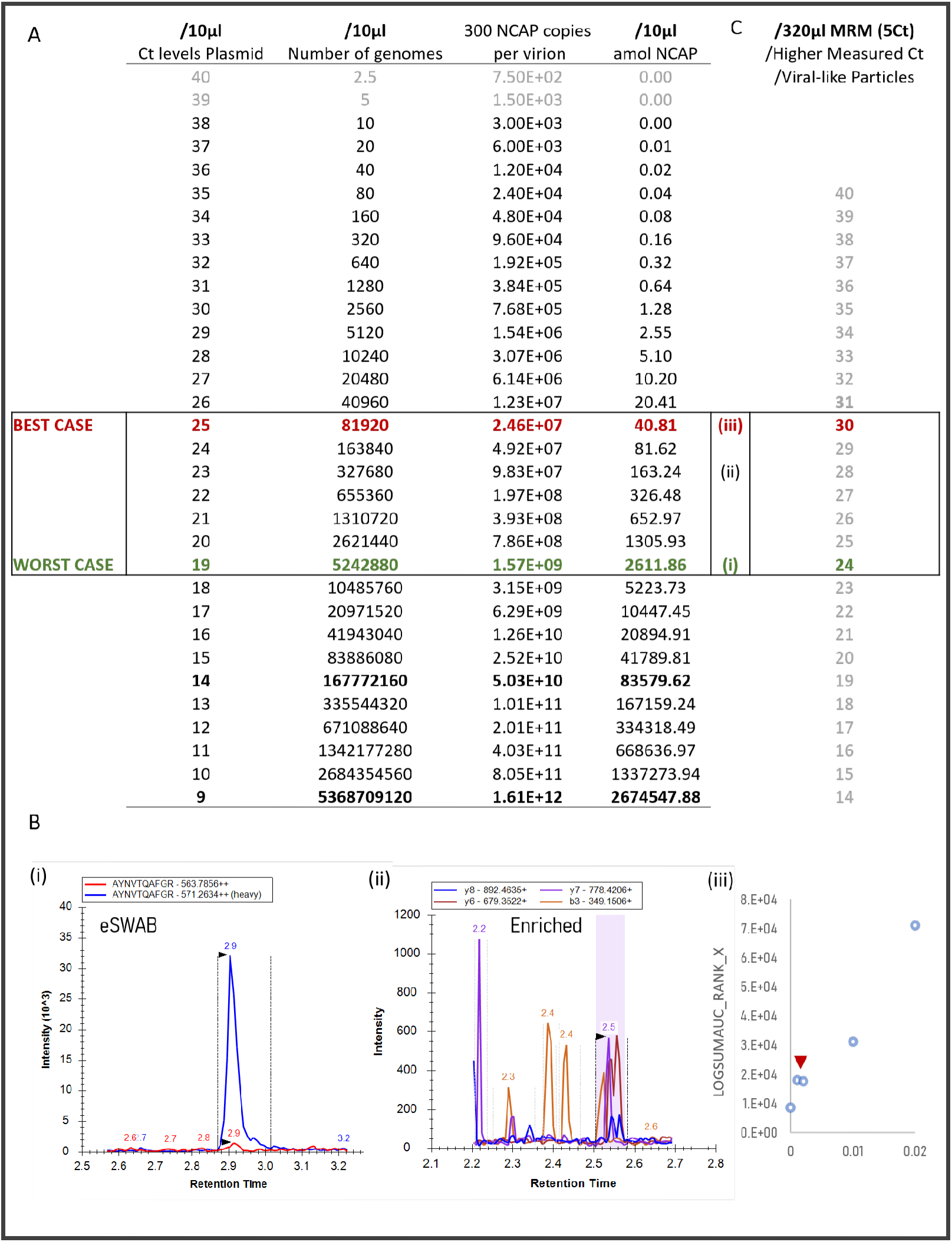
A projection of MRM capabilities. **(A) Absolute quantities theoretically correlate Ct to the MRM assay**. 10 genomes present in 10µl are detected by a Ct of 38 in perfect conditions (i.e. plasmids) in our in-house RT-PCR. Assuming 300 NCAP molecules per virion, a direct translation into amoles is possible. The highlighted portion of the table depicts **(B) three different limits of detection for the MRM assay**. (i) AYNVTQAFGR With the Cov-MS QConCAT in eSwab background, (ii) following peptide enrichment (signal taken from in solution dilution from **figure 1D**) and (iii) possibly by more data-driven approaches or summing all signal at the elution window of peptides without peak integration. **(C) The potential practical correlation between Ct and MRM assay**. By loading up to 32 times more by enrichment of targets could yield another 5Ct values of sensitivity. This is enforced by the fact that clinical RT-PCRs are expected to be higher than on plasmids and the MRM signal could be higher because of VLPs.

The Cov-MS labs have shown that the latest generation tandem quadrupole instruments are sensitive enough to detect patients passed a Ct 20 in medium. In eSwab, an LOD of NCAP of at least 0.1 ng oc was attained when the Cov-MS QConCAT was used for peak picking on the latest generation instruments, i.e. a 7500 from Sciex (**Supplementary Data 21**), in line with our earlier LODs depicted in **Figure 1D**. However, moving beyond Ct values of 25 – as shown before on high resolution platforms (40,43) – will thus require peptide enrichment. Protein or peptide immuno-enrichment can be done either through antibodies (e.g. SISCAPA) or aptamers (44,45). This way, the few molecules that are present in a sample can be measured almost perfectly, i.e. in isolation with minimal interference. In the process, this will open the way to other COVID-19 sampling matrices, like gargle solution, saliva, and plasma and finally alleviate the pressure from ever-changing media being used in the clinic. Together with the Cov-MS SIL standard that ascertains the correct detection of low signals and a low coefficient of variation, this could lead to a an automatable, robust and sensitive alternative for RT-PCR.

Will this be enough to become clinically relevant? Well, the limit of contagiousness was previously estimated be close to Ct 33-34 (46). Additionally, one particularly interesting application of the MRM assay would be the second phase screening of pooled patient PCR assays. More specifically, it was recently suggested that for Belgium a population-wide weekly screening could be possible if patients are pooled by 32 (47). This is the equivalent of lowering sensitivity by 5Ct values and this implies that the final MRM assay could be perfectly suited to screen the individual patients in positive batches.

#### Increasing Throughput

A major drawback of the current LC-MS based workflow is that data acquisition cannot be parallelized on a single instrument, contrary to RT-PCR assays that use multi-well plates. Yet, using 8-minute gradients, one instrument can already measure over 150 patients/day and by multiplexing LC pumps this can be increased to ∼500 per day (27). One member of the consortium (Alderley Analytical) additionally looked at the application of two-minute gradients and showed that these should be feasible when the matrix is relatively clean (**Supplementary data 15**), increasing the throughput to 600 patients per instrument per day without parallelization. With prior immuno-enrichment, gradients as low as 1 minute are within reach. At least from the sample preparation perspective, there is no reason why robotics cannot be deployed, as is the case for RT-PCR preparation.

One disruptive recent technology to further increase throughput is acoustic MS. In the implementation from Sciex (EchoMS), nanoliter droplets are acoustically ejected from the sample and into the instrument at a rate of 3Hz, with each droplet measuring up to five transitions (48). In time, this could lead to sampling an estimated 30.000 patients a day, per instrument. Likewise MALDI-ToF instruments could provide very high throughput if they could be shown to have the required sensitivity following immuno-enrichment of target peptides.

#### Screening multiple pathogens in a single assay

Using the proposed workflow, it is relatively straightforward to add mutations or even other pathogens for screening in a single assay. These include other corona viruses as well as e.g. influenza. This will greatly increase the impact of protein detection by MRM in the clinic.

#### Making Data Analysis Clinically applicable

To cope with all the resulting data, the automation of data analysis in a clinically applicable way will still require some efforts. However, with recent advances in machine learning in the proteomics field, we are optimistic that a community effort in data analysis can move forward very quickly (49). This includes automated diagnosis based on the MRM data. Initiatives such as EuBIC, prominent advocates of data sharing, provide the perfect basis for seeding such efforts. Meeting diagnostic assay criteria required by regulatory agencies, including calibrators/internal standards, sample preparation, instrumentation, as well as data review and reporting, all mentioned and detailed here, requires consideration from a broader deployment perspective. Fortunately, however, these requirements are understood, as previous efforts such as the National Cancer Institute’s Clinical Proteomic Technologies for Cancer initiative and the HUPO Proteomics Standards Initiative have aided the acceptance and validation processes by introducing the “verification” concept using MRM technologies, similarly to the **Accreditation Standards** concept and results introduced in this manuscript. Together, this illustrates that collaboration will be key to the final success of a “fit-for-purpose” MRM assay (50).

**In conclusion, we describe the full pipeline for developing an MS-based assay for viral presence directly on clinical samples and with conventional instrumentation. The current assay allows detection of up to 200 patients below Ct 25 per instrument per day. By removing matrix, 1 minute per patient could result in up to nearly 1500 patients of potentially up to a Ct 30 on a single LC-MS setup. It is also clear that the free sharing of all acquired data will be essential in such an endeavor. At the same time, we cannot know for sure when this protein-based mass spectrometry-based test will be ready for the clinic or when the clinic will be ready for a protein-based mass spectrometry test. Nevertheless, it is clear that mass spectrometry stands a chance of matching PCR’s output and economies of scale, if a collective effort by the community is applied. The UK is currently setting the scene, building on collaborative efforts like COVID-MSC (23) and Cov-MS and others will most likely follow. And what is certain, is that not trying would be scientifically negligent given the critical importance of the issue**.

## Supporting information

Detailed Methods Section

## Data Availability

The LC-MS data generated within the Cov-MS consortium is shared and browsable through Panorama Public (https://panoramaweb.org/CovMS.url) and DDA, predicted and chromatogram libraries as well as the narrow window DIA data have been deposited to the ProteomeXchange Consortium via the PRIDE partner repository (24) with the dataset identifier PXD022550 and 10.6019/PXD022550.

## Acknowledgements

This research was funded by grants from the Research Foundation Flanders (FWO): B.V.P (grant number 11B4518N), R.G. (grant number 1S50918N), T.V.D.B (grant number 1S90918N), P.R, L.M (grant number G032816N), L.M. (grant number G042518N), N.D. (1S21017N), and M.D. (12E9716N); by the BOF-COVID-19 project grant (01C01920) from Ghent University; and by the European Union’s Horizon 2020 Programme under Grant Agreement 823839 (H2020-INFRAIA-2018-1). Ethical approval: Obtained from University Hospital Leuven Ethics Committee with n° S63879.

## Data availability

The mass spectrometry datasets generated during the current study are being shared through the Skyline Panorama Public interface for easy accessibility (https://panoramaweb.org/CovMS.url) and are being deposited to the ProteomeXchange Consortium via the PRIDE partner repository with dataset identifier: PXD022550. The supplementary material will be made available through the Microsoft Teams group: “Cov-MS Digital Incubator”. Please send an email to to obtain access. The detailed methods section is still being revised.

## Code availability

MS^2^PIP, DeepLC, EncyclopeDIA and the Machine Learning algorithms are open source, licensed under the Apache-2.0 License, and are hosted on https://github.com/compomics/ms2pip_c, https://github.com/compomics/DeepLC, https://bitbucket.org/searleb/encyclopedia/wiki/Home and https://github.com/compomics/Cov-MS-scoring.

## References

1. OECD. Testing for COVID-19: A way to lift confinement restrictions. 2020 [cited 2020 Aug 19];(May):1–21. Available from: https://read.oecd-ilibrary.org/view/?ref=129_129658-l62d7lr66u&title=Testing-for-COVID-19-A-way-to-lift-confinement-restrictions

2. Surkova E, Nikolayevskyy V, Drobniewski F. False-positive COVID-19 results: hidden problems and costs. Lancet Respir Med [Internet]. 2020 Sep [cited 2020 Oct 14];0(0). Available from: https://doi.org/10.1016/S2213-2600

3. SARS-CoV-2 Reference Panel Comparative Data | FDA [Internet]. [cited 2020 Oct 14]. Available from: https://www.fda.gov/medical-devices/coronavirus-covid-19-and-medical-devices/sars-cov-2-reference-panel-comparative-data

4. ASM. ASM Expresses Concern about Coronavirus Test Reagent Shortages [Internet]. 2020 [cited 2020 Aug 19]. Available from: https://asm.org/Articles/Policy/2020/March/ASM-Expresses-Concern-about-Test-Reagent-Shortages

5. Are you infectious if you have a positive PCR test result for COVID-19? - CEBM [Internet]. [cited 2020 Oct 14]. Available from: https://www.cebm.net/covid-19/infectious-positive-pcr-test-result-covid-19/

6. Atkinson B, Petersen E. SARS-CoV-2 shedding and infectivity [Internet]. Vol. 395, The Lancet. Lancet Publishing Group; 2020 [cited 2020 Oct 14]. p. 1339–40. Available from: http://ees.elsevier.com/thelancet/www.thelancet.com

7. Nachtigall FM, Pereira A, Trofymchuk OS, Santos LS. Detection of SARS-CoV-2 in nasal swabs using MALDI-MS. Nat Biotechnol. 2020;

8. Mak GC, Cheng PK, Lau SS, Wong KK, Lau CS, Lam ET, et al. Evaluation of rapid antigen test for detection of SARS-CoV-2 virus. J Clin Virol. 2020;

9. Ihling C, Tänzler D, Hagemann S, Kehlen A, Hüttelmaier S, Sinz A. Mass Spectrometric Identification of SARS-CoV-2 Proteins from Gargle Solution Samples of COVID-19 Patients. bioRxiv [Internet]. 2020 Jan 1;2020.04.18.047878. Available from: http://biorxiv.org/content/early/2020/04/19/2020.04.18.047878.abstract

10. Bezstarosti K, Lamers MM, Haagmans BL, Demmers JAA. TARGETED PROTEOMICS FOR THE DETECTION OF SARS-COV-2 PROTEINS. bioRxiv [Internet]. 2020 Jan 1;2020.04.23.057810. Available from: http://biorxiv.org/content/early/2020/04/23/2020.04.23.057810.abstract

11. Giri R, Bhardwaj T, Shegane M, Gehi BR, Kumar P, Gadhave K, et al. When Darkness Becomes a Ray of Light in the Dark Times: Understanding the COVID-19 via the Comparative Analysis of the Dark Proteomes of SARS-CoV-2, Human SARS and Bat SARS-Like Coronaviruses. bioRxiv [Internet]. 2020 Jan 1;2020.03.13.990598. Available from: http://biorxiv.org/content/early/2020/04/03/2020.03.13.990598.abstract

12. Grenga L, Gallais F, Pible O, Gaillard J-C, Gouveia D, Batina H, et al. Shotgun proteomics of SARS-CoV-2 infected cells and its application to the optimisation of whole viral particle antigen production for vaccines. bioRxiv [Internet]. 2020 Jan 1;2020.04.17.046193. Available from: http://biorxiv.org/content/early/2020/04/17/2020.04.17.046193.abstract

13. Ortea I, Bock J-O. Re-analysis of SARS-CoV-2 infected host cell proteomics time-course data by impact pathway analysis and network analysis. A potential link with inflammatory response. bioRxiv [Internet]. 2020 Jan 1;2020.03.26.009605. Available from: http://biorxiv.org/content/early/2020/03/28/2020.03.26.009605.abstract

14. Gordon DE, Jang GM, Bouhaddou M, Xu J, Obernier K, O’Meara MJ, et al. A SARS-CoV-2-Human Protein-Protein Interaction Map Reveals Drug Targets and Potential Drug-Repurposing. bioRxiv [Internet]. 2020 Jan 1;2020.03.22.002386. Available from: http://biorxiv.org/content/early/2020/03/27/2020.03.22.002386.abstract

15. Orsburn BC, Jenkins C, Miller SM, Neely BA, Bumpus NN. *In silico* approach toward the identification of unique peptides from viral protein infection: Application to COVID-19. bioRxiv [Internet]. 2020 Jan 1;2020.03.08.980383. Available from: http://biorxiv.org/content/early/2020/04/10/2020.03.08.980383.abstract

16. Davidson AD, Williamson MK, Lewis S, Shoemark D, Carroll MW, Heesom K, et al. Characterisation of the transcriptome and proteome of SARS-CoV-2 using direct RNA sequencing and tandem mass spectrometry reveals evidence for a cell passage induced in-frame deletion in the spike glycoprotein that removes the furin-like cleavage site. bioRxiv [Internet]. 2020 Jan 1;2020.03.22.002204. Available from: http://biorxiv.org/content/early/2020/03/24/2020.03.22.002204.abstract

17. Grossegesse M, Hartkopf F, Nitsche A, Schaade L, Doellinger J, Muth T. Perspective on Proteomics for Virus Detection in Clinical Samples. J Proteome Res [Internet]. 2020 Oct 22 [cited 2020 Oct 23];acs.jproteome.0c00674. Available from: https://pubs.acs.org/doi/10.1021/acs.jproteome.0c00674

18. Neuman BW, Buchmeier MJ. Supramolecular Architecture of the Coronavirus Particle. In: Advances in Virus Research. Academic Press Inc.; 2016. p. 1–27.

19. Swann H, Sharma A, Preece B, Peterson A, Eldridge C, Belnap D, et al. Minimal system for assembly of SARS-CoV-2 virus like particles. bioRxiv [Internet]. 2020 Jul 15 [cited 2020 Aug 19];2020.06.01.128058. Available from: https://doi.org/10.1101/2020.06.01.128058

20. Plante JA, Liu Y, Liu J, Xia H, Johnson BA, Lokugamage KG, et al. Spike mutation D614G alters SARS-CoV-2 fitness. Nature [Internet]. 2020 Oct 26 [cited 2020 Nov 4];1–9. Available from: http://www.nature.com/articles/s41586-020-2895-3

21. Bojkova D, Klann K, Koch B, Widera M, Krause D, Ciesek S, et al. SARS-CoV-2 infected host cell proteomics reveal potential therapy targets. 2020 Mar 11 [cited 2020 Aug 20]; Available from: https://www.researchsquare.com/article/rs-17218/v1

22. Pino LK, Searle BC, Bollinger JG, Nunn B, MacLean B, MacCoss MJ. The Skyline ecosystem: Informatics for quantitative mass spectrometry proteomics [Internet]. Vol. 39, Mass Spectrometry Reviews. John Wiley and Sons Inc.; 2020 [cited 2020 Nov 12]. p. 229–44. Available from: https://pubmed.ncbi.nlm.nih.gov/28691345/

23. Struwe W, Emmott E, Bailey M, Sharon M, Sinz A, Corrales FJ, et al. The COVID-19 MS Coalition—accelerating diagnostics, prognostics, and treatment [Internet]. Vol. 395, The Lancet. Lancet Publishing Group; 2020 [cited 2020 Nov 6]. p. 1761–2. Available from: https://www.nouveal.com/

24. Perez-Riverol Y, Csordas A, Bai J, Bernal-Llinares M, Hewapathirana S, Kundu DJ, et al. The PRIDE database and related tools and resources in 2019: Improving support for quantification data. Nucleic Acids Res. 2019;

25. Van Puyvelde B, Willems S, Gabriels R, Daled S, De Clerck L, Vande Casteele S, et al. Removing the Hidden Data Dependency of DIA with Predicted Spectral Libraries. Proteomics [Internet]. 2020 Feb 5;20(3–4):e1900306. Available from: http://www.ncbi.nlm.nih.gov/pubmed/31981311

26. Laboratory biosafety guidance related to coronavirus disease (COVID-19) [Internet]. [cited 2020 Nov 4]. Available from: https://www.who.int/publications/i/item/laboratory-biosafety-guidance-related-to-coronavirus-disease-(covid-19)

27. Cardozo KHM, Lebkuchen A, Okai GG, Schuch RA, Viana LG, Olive AN, et al. Fast and low-cost detection of SARS-CoV-2 peptides by tandem mass spectrometry in clinical samples. 2020 May 17 [cited 2020 Nov 3]; Available from: https://www.researchsquare.com/article/rs-28883/v1

28. Zecha J, Lee C-Y, Bayer FP, Meng C, Grass V, Zerweck J, et al. Data, Reagents, Assays and Merits of Proteomics for SARS-CoV-2 Research and Testing. 2020 [cited 2020 Nov 3]; Available from: https://doi.org/10.1074/mcp.RA120.002164

29. Gouveia D, Grenga L, Gaillard J, Gallais F, Bellanger L, Pible O, et al. Shortlisting SARS-CoV-2 Peptides for Targeted Studies from Experimental Data-Dependent Acquisition Tandem Mass Spectrometry Data. Proteomics [Internet]. 2020 Jul 21 [cited 2020 Nov 4];20(14):2000107. Available from: https://onlinelibrary.wiley.com/doi/abs/10.1002/pmic.202000107

30. Van Puyvelde B, Willems S, Gabriels R, Daled S, De Clerck L, Vande Casteele S, et al. Removing the Hidden Data Dependency of DIA with Predicted Spectral Libraries. Proteomics [Internet]. 2020 Feb 5;20(i3–4):e1900306. Available from: http://www.ncbi.nlm.nih.gov/pubmed/31981311

31. Panchaud A, Scherl A, Shaffer SA, von Haller PD, Kulasekara HD, Miller SI, et al. Precursor acquisition independent from ion count: how to dive deeper into the proteomics ocean. Anal Chem [Internet]. 2009 Aug 1;81(15):6481–8. Available from: https://pubs.acs.org/doi/10.1021/ac900888s

32. Gabriels R, Martens L, Degroeve S. Updated MS^2^PIP web server delivers fast and accurate MS^2^ peak intensity prediction for multiple fragmentation methods, instruments and labeling techniques. Nucleic Acids Res [Internet]. 2019 Jul 2;47(W1):W295–9. Available from: https://academic.oup.com/nar/article/47/W1/W295/5480903

33. Bouwmeester R, Gabriels R, Hulstaert N, Martens L, Degroeve S, †vib-Ugent ‡. DeepLC can predict retention times for peptides that carry as-yet unseen modifications. bioRxiv [Internet]. 2020 Mar 29 [cited 2020 Oct 14];2020.03.28.013003. Available from: https://doi.org/10.1101/2020.03.28.013003

34. Searle BC, Pino LK, Egertson JD, Ting YS, Lawrence RT, MacLean BX, et al. Chromatogram libraries improve peptide detection and quantification by data independent acquisition mass spectrometry. Nat Commun [Internet]. 2018 Dec 3;9(1):5128. Available from: http://www.nature.com/articles/s41467-018-07454-w

35. Šlechtová T, Gilar M, Kalíková K, Tesařová E. Insight into Trypsin Miscleavage: Comparison of Kinetic Constants of Problematic Peptide Sequences. Anal Chem. 2015;

36. Yao H, Song Y, Chen Y, Wu N, Xu J, Sun C, et al. Molecular Architecture of the SARS-CoV-2 Virus. Cell. 2020;

37. Maarten Dhaenens. Cov-MS Consortium Launch (Teams Meeting) [Internet]. 2020 [cited 2020 Aug 21]. Available from: http://genesis.ugent.be/uvpublicdata/Cov-MS_launch.mp4

38. Comprehending COVID-19: Maximizing LC-MS Detection Dynamic Range for Multiple Reaction Monitoring Based SARS-CoV-2 Analysis: Waters [Internet]. [cited 2020 Oct 20]. Available from: https://www.waters.com/nextgen/us/en/library/application-notes/2020/comprehending-covid-19-maximizing-lc-ms-detection-dynamic-range-for-multiple-reaction-monitoring-based-sars-cov-2-analysis.html

39. Comprehending COVID-19: Multiple Reaction Monitoring Transition Selection and Optimization Strategies for LC-MS Based SARS-CoV-2 Detection : Waters [Internet]. [cited 2020 Oct 20]. Available from: https://www.waters.com/nextgen/us/en/library/application-notes/2020/comprehending-covid-19-multiple-reaction-monitoring-transition-selection-and-optimization-strategies-for-lc-ms-based-sars-cov-2-detection.html

40. Gouveia D, Miotello G, Gallais F, Gaillard J-C, Debroas S, Bellanger L, et al. Proteotyping SARS-CoV-2 Virus from Nasopharyngeal Swabs: A Proof-of-Concept Focused on a 3 Min Mass Spectrometry Window. J Proteome Res [Internet]. 2020 Aug 5 [cited 2020 Nov 3]; Available from: https://dx.doi.org/10.1021/acs.jproteome.0c00535

41. Beynon RJ, Doherty MK, Pratt JM, Gaskell SJ. Multiplexed absolute quantification in proteomics using artificial QCAT proteins of concatenated signature peptides. Nat Methods [Internet]. 2005 Aug [cited 2020 Oct 23];2(8):587–9. Available from: https://pubmed.ncbi.nlm.nih.gov/16094383/

42. Peaks of light and heavy peptides have unequal peak boundaries: /home/support [Internet]. [cited 2020 Oct 26]. Available from: https://skyline.ms/announcements/home/support/thread.view?rowId=48979

43. Nikolaev EN, Indeykina MI, Brzhozovskiy AG, Bugrova AE, Kononikhin AS, Starodubtseva NL, et al. Mass-Spectrometric Detection of SARS-CoV-2 Virus in Scrapings of the Epithelium of the Nasopharynx of Infected Patients via Nucleocapsid N Protein. [cited 2020 Nov 3]; Available from: https://dx.doi.org/10.1021/acs.jproteome.0c00412

44. Zhang L, Fang X, Liu X, Ou H, Zhang H, Wang J, et al. Discovery of sandwich type COVID-19 nucleocapsid protein DNA aptamers. Chem Commun. 2020;

45. Razavi M, Leigh Anderson N, Pope ME, Yip R, Pearson TW. High precision quantification of human plasma proteins using the automated SISCAPA Immuno-MS workflow. N Biotechnol. 2016;

46. La Scola B, Le Bideau M, Andreani J, Hoang VT, Grimaldier C, Colson P, et al. Viral RNA load as determined by cell culture as a management tool for discharge of SARS-CoV-2 patients from infectious disease wards. Eur J Clin Microbiol Infect Dis [Internet]. 2020 Jun 1 [cited 2020 Nov 3];39(6):1059–61. Available from: https://doi.org/10.1007/s10096-020-03913-9

47. Libin P, Willem L, Verstraeten T, Torneri A, Vanderlocht J, Hens N. Assessing the feasibility and effectiveness of household-pooled universal testing to control COVID-19 epidemics. medRxiv [Internet]. 2020 Oct 6 [cited 2020 Nov 12];2020.10.03.20205765. Available from: https://doi.org/10.1101/2020.10.03.20205765

48. Echo MS | SCIEX [Internet]. [cited 2020 Oct 29]. Available from: https://sciex.com/products/integrated-solutions/echo-ms?utm_source=adwords+search&tm_medium=cpc&tm_campaign=2020+product+launches&utm_term=g-b-h-a-echo+ms+search+ads-may-20&tm_content=echo+ms&clid=CjwKCAjw0On8BRAgEiwAincsHFdmzI5srCuEBblfHuixR30D0_rYS7mJiXTCo8bVxiYi0ZGHSwJZCBoCkdQQAvD_BwE

49. Bouwmeester R, Gabriels R, Van Den Bossche T, Martens L, Degroeve S. The Age of Data-Driven Proteomics: How Machine Learning Enables Novel Workflows. Proteomics. 2020;

50. Carr SA, Abbatiello SE, Ackermann BL, Borchers C, Domon B, Deutsch EW, et al. Targeted peptide measurements in biology and medicine: Best practices for mass spectrometry-based assay development using a fit-for-purpose approach. Mol Cell Proteomics [Internet]. 2014 Mar 1 [cited 2020 Nov 4];13(3):907–17. Available from: http://www.mcponline.org

