## Supplementary material for "Cov-MS: a community-based template assay for clinical MS-based protein detection in SARS-CoV-2 patients": Detailed Methods Section

#### Discovery Phase (Blue)

##### Sample Preparation

50µl of a patient sample (1/60) was precipitated by adding 450µl (9 volumes) of ice-cold acetone (-20°C). After spinning at 16.000g and 0°C, the supernatant was discarded and 1µg of Trypsin/Lys-C mix (Promega) in 60µl 500 mM triethylammonium bicarbonate (TEABC) buffer was added. This was followed by an incubation step of four hours at 37°C, to facilitate trypsin digestion. Next, 20µl of this sample was prepared for analysis in a final concentration of 0.1% formic acid (FA) of which 2µl was injected into the LC-MS system. The protocol was validated on a dilution series of two recombinant Covid-19 proteins NCAP_SARS2 and SPIKE_SARS2 (Sino Biological, Beijing, China) which were found to be most abundant in public data on SARS-COV 2. A triplicate dilution series of 250 µL of negative patient UTM medium was spiked with different amounts of recombinant protein (500, 100, 50 ,10, 5, 1, 0.5, 0.1, 0.05, 0ng) resulting in a concentration range of 1ng-300fg on column.

Note: After receiving some questions concerning the concentration of the TEABC buffer, we evaluated the digest efficiency with a less concentrated TEABC solution (50mM) and with 50 mM of ammonium bicarbonate (ABC). Both performed comparable to the 500mM TEABC solution (data not shown).

##### Data Acquisition

###### *Data Dependent Acquisition (DDA)*

DDA was performed with reversed phase LC-MS using an Eksigent NanoLC 425 (Eksigent, Dublin, CA) system operated in microflow mode and coupled to a TripleTOF5600 and a TripleTOF6600+ mass spectrometer (AB Sciex, Concord, Ontario,Canada). The LC system was operated with 0.1% Formic Acid (FA) in water (Buffer A) and 0.1% FA in ACN (Buffer B). Peptides were trapped on a C18 trap column (YMC) at 10µL/min for 3 minutes and separated on a Phenomenex Luna Omega Polar C18 (150 x 0.3 mm) column at 5 µL/min. A 60 min LC-gradient from 3-55% B was followed by a washing and equilibration step before proceeding to the next injection. MS1 spectra were collected from 400-1250 m/z for 250 ms. The 30 most intense precursor ions with charge state 2-4 that exceeded 300 counts per second were selected for fragmentation, and the corresponding fragmentation MS2 spectra were collected between 100-1500 m/z for 50 ms. After fragmentation, precursor ions were dynamically excluded from reselection for 10 s. Rolling collision energy with a collision energy spread of 0V was used for fragmentation to mimic SWATH like fragmentation. The TripleTOF5600 was equipped with a 50µm DuoSpray Ion source, while the 6600+ was equipped with an Optiflow Ion source.

###### *Sequential Window Acquisition of all Theoretical fragment ion spectra (SWATH)*

SWATH data were collected with the same TripleTOF6600+ system as described above, with identical chromatographic conditions and using a 64 variable acquisition window scheme, optimized by Navarro et al [1]. The SWATH MS2 spectra were collected in high-sensitivity mode from 100 to 1500 m/z, for 50 ms. Before each SWATH MS cycle an additional MS1 survey scan in high sensitivity mode was recorded for 150 ms, resulting in a total duty cycle of ~3.4 s.

###### *Narrow-Window DIA*

To build the chromatogram library, the SCIEX6600+ was configured to acquire eight gas phase fractionated injections of 100 m/z (400-500,500-600,...) each acquired with overlapping 4Da windows, as described by Searle et al. [2]. To reduce the acquisition time, we applied a 10min LC gradient (3-30% ACN), this kept the total duty cycle manageable to 1.45 seconds and with an accumulation time of 100 and 25 ms respectively, for the MS1 and MS2 spectra.

##### *MRM optimization*

For each peptide, the cone and collision energies were optimized within 8-minute runs as described in the SOP (**Supplementary data 8**). This can be done by using 5-7 MRM transitions in one run varying in the second/third digit of the parent/daughter, each measured with a different cone/fixed collision or the other way around. More specifically, using a 1 ng/µL stock solution of digested recombinant NCAP_SARS2 and SPIKE_SARS2 proteins, the retention time of each peptide was measured using 17 individual MRM transitions. In parallel, the cone parameters were optimized, resulting in 17 dedicated MRM-files. The collision energy for each peptide, and each of the 5 candidate fragments (using the optimal cone energy) was optimized next, resulting in 17 * 5 dedicated MRM-files. Next, a scheduled MRM-file was created, retaining the most sensitive peptides based on human inspection. All our MRM optimization was performed on a Waters Acquity LC system coupled to a Xevo TQ-S (Waters, Winslow). The UTM dilution series and the 20-patient sample were freshly prepared, as described before, and acquired with the optimized MRM method as described in the SOP (Supplementary data 8). The injection volume for the MRM method was 5 µL (2,5x higher load on column when compared to the DDA/SWATH data). The data was analyzed in Skyline Daily using a template file containing the 17 target peptides. Peak boundaries were manually adjusted, as this was required, considering the amount of interfering transitions, originating from the matrix. Note that settings these peak boundaries could be considered as a subjective task which might introduce bias in the data analysis.

#### Data-Analysis

##### *DDA (MS-GF+ / Mascot Daemon)*

*T*he data was peak picked with MSConvert (Version 3.0.20070) at the MS1 level using the built-in vendor specific algorithms. The peak lists (.mgf files) obtained from MS/MS spectra were identified using MS-GF+ (version v2018.04.09) [3]. The searches were conducted using SearchGUI version 3.3.17 [4]. Protein identification was conducted against a concatenated target/decoy database of Homo sapiens reference proteome, SARS-Cov-2 database, and the cRAP database of contaminants (https://thegpm.org/cRAP) (downloaded from Uniprot on the 4th of April 2020). The decoy sequences were created by reversing the target sequences in SearchGUI, and the identification settings were as follows: specific cleavage with trypsin with a maximum of two missed cleavages; peptide charges from 2+ to 4+; 10.0 ppm as MS1 tolerance and 0.02 Da as MS2 tolerance; Oxidation of M, Oxidation of P and Acetylation of protein N-termini as variable modification. Peptides and proteins were inferred from the spectrum identification results using PeptideShaker version 1.16.43 [5] PSMs, peptides and proteins were validated at a 1% FDR estimated using the decoy hit distribution. The peptides were exported to the Unipept web application for an explorative taxonomic analysis [6,7]. Additional searches were performed with Mascot v2.7.0 using the same concatenated database as described above. Following search parameters were applied: trypsin as digestion enzyme, a maximum of two missed cleavages, peptide charges 2+ to 4+, peptide mass tolerance of 10 ppm, fragment ion tolerance of 50 ppm and oxidation of Methionine and Deamidation (NQ) as variable modification.

##### *Narrow-Window DIA (EncyclopeDIA)*

A FASTA protein database was compiled by concatenating the protein identifications obtained with Mascot for the 20 patient samples with the SARS-CoV-2 proteome. Using the fasta2speclib script (as part of the MS²PIP Python package, version 3.6.1) [8], a spectral library containing predicted spectra (MS²PIP) and predicted retention times (DeepLC) [9] was generated for all possible peptide-charge-modification combinations, given the proteins in the FASTA input. The MSP output file was converted to a .dlib file, using the conversion tools embedded in EncyclopeDIA (Supplementary Data 1B). The EncyclopeDIA GUI, version 0.9.0, was downloaded from bitbucket (https://bitbucket.org/searleb/EncyclopeDIA) and run on a Microsoft Windows 10-based system (Lenovo Thinkstation, Intel Xeon E5-2620, 24 processors, 128 GB ram).

The eight gas phase fractions (GP) of 100 m/z each covering a 400-1200 m/z range, were peak picked and demultiplexed into 2m/z (narrow window DIA) windows and finally converted into .mzML's by MSConvert with following parameters:

> **Peak picking:** Vendor specific algorithms (algorithms available for all vendors, except Waters)
> **Demultiplexing:** overlap only with a mass error of 10 ppm

The predicted spectral library was set as library to search the narrow window DIA data. Additional settings are described below, in the exact order as required by the EncyclopeDIA GUI.

> **Background:** 200406_Human_Covid19.fasta
> **Target/Decoy approach:** Normal Target/Decoy
> **Data Acquisition Type:** Non-Overlapping DIA
> **Enzyme:** Trypsin
> **Fragmentation:** CID/HCD (b- and y- fragments)
> **Precursor/Fragment/Library Mass Tolerance:** 10.0 ppm
> **Percolator Version:** v3-01
> **Number of Quantitative Ions:** 5
> **Minimum number of Quantitative Ions:** 3
> **Number of Cores:** 24

Finally, the peptide and protein identification results were exported as .BLIB file to facilitate peptide-centric analysis of the SWATH data.

##### *SWATH (Skyline Daily)*

The SWATH data from the UTM dilution series and the 20 patient samples were analyzed using the Skyline-daily (version 20.1.9.234) software developed by the MacCoss Lab. Only tryptic peptides with precursor charge state 2+ and 3+ and fragment ion charges 1+ and 2+ were considered. The 10-best b- and y-ions for each precursor from a library spectrum were retained, and a minimum of 3 product ions was required to be included in the target list. Oxidation (M) and Deamidation (NQ) were considered as variable modifications. MS1 and MS2 filtering were performed with "TOF mass analyzer" set and with a resolving power of 30k. The 64 variable window isolation scheme was generated by importing one of the SWATH .wiff files. Finally, an iRT calculator was created by manual selecting 14 peptides, nicely spread over the LC gradient, related to albumin which is highly abundant in UTM medium. Retention time filtering was applied by only using scans within 5 minutes of the predicted retention time. Reversed sequence decoys were added to the target list, to enable mProphet training. Peak integration boundaries were reviewed and manually adjusted before exporting a report (.csv) containing peptide sequence, BioReplicate and Area amongst others. The library dot product, together with the correlation between spiked concentration and peak area were assessed for all peptides related to NCAP_SARS2 and SPIKE_SARS2. From these results, we were able to correctly classify 18/20 patient samples and additionally, we were able to identify 17 responsive target peptides (**Supplementary Data 4**). These 17 peptides with their corresponding fragment ion ranks were reported in the SOP to enable translation of the discovery data into a targeted MRM assay

##### *Evolutionary conservation and taxonomic analysis methods*

The 17 selected candidate peptides were subjected to a taxonomic analysis using the Unipept web application (version 4.3) [6]. The UniProtKB version present in Unipept 4.3 is the 2020-01 release, which did not include novel SARS-Cov-2 proteins. In Unipept, the "Equate I/L" option was enabled.

Each peptide could be categorized in four possible categories. In the first category, the lowest common ancestor (LCA) was assigned to root, which means that this peptide could be present in many different organisms. This was the case for two peptides (DQVILLNK and LNQLESK). In the additional analysis with Unipept CLI 2.0 [10], all taxa were retrieved. Here we could observe that these two peptides are part of many other organisms (Unipept_taxa.txt). DQVILLNK was found 10 times in Unipept, of which two times in Coronaviridae, and eight times in unrelated species. LNQLESK was found 100 times in Coronaviridae of the 177 hits in total. In the second category, the LCA was assigned to the Coronaviridae family. Here, the peptides are not uniquely present in SARS-Cov-2 but could also be found in other members of this family. In our analysis, six peptides fall in this category (AYNVTQAFGR, GQQQQGQTVTK, HWPQIAQFAPSASAFFGMSR, RGPEQTQGNFGDQELIR, RSFIEDLLFNK, and SFIEDLLFNK). In the third category, one peptide (NPANNAAIVLQLPQGTTLPK) was uniquely assigned to the species Severe acute respiratory syndrome-related coronavirus, also known as SARS-Cov. This peptide is therefore not uniquely to SARS-Cov-2 but is more specific than being assigned to multiple members of the Coronaviridae family. The fourth category consists of eight peptides (ADETQALPQR, DGIIWVATEGALNTPK, EDLKFPR, GQGVPINTNSSPDDQIGYYR, GWIFGTTLDSK, IGMEVTPSGTWLTYTGAIK, KQQTVTLLPAADLDDFSK, and MAGNGGDAALALLLLDR) that are not found in Unipept (and thus not in the Uniprot 2020-01 release). These peptides map therefore uniquely back to Severe acute respiratory syndrome-related coronavirus 2 (SARS-Cov-2).

The selected peptides were then mapped onto the three-dimensional (3D) protein structures obtained from SARS-Cov-2 dedicated page from RCSB-PDB [11]. 12 out of 17 peptides were mapped onto SPIKE_SARS2 (SFIEDLLFNK, RSFIEDLLFNK, GWIFGTTLDSK), NCAP_SARS2 RNA binding domain (DGIIWVATEGALNTPK, EDLKFPR, GQGVPINTNSSPDDQIGYYR, NPANNAAIVLQLPQGTTLPK) and NCAP_SARS2 C-terminal dimerization domain (AYNVTQAFGR, DQVILLNK, HWPQIAQFAPSASAFFGMSR, IGMEVTPSGTWLTYTGAIK, RGPEQTQGNFGDQELIR). Mapped peptides are highlighted in structures (Figure 2B). In-order to perform the evolutionary sequence conservation on protein sequences and map them onto the protein structures, we obtained all the protein sequences of coronoviridae family from UniProt-KB. The conservation mapping was done with Scop3D [12]using the respective coronoviridae protein sequences and structure for that particular protein (for example to map the conservation on 6VXX structure, only spike glycoprotein sequences from coronoviridae family were used). The conservation scores were colored red (0% conserved) to blue (100% conserved) scale (Figure 2B). The images were generated using the PyMOL Molecular Graphics System, version 2.3.4 [13].

### The Cov-MS consortium (Red)

#### Sample Preparation optimization

##### *Optimizing digest efficiency*

Different sample preparation optimization protocols were performed in quintuplicate on 20ng SPIKE_SARS2 and NCAP_SARS2 in 100µL of UTM- or eSwab medium from a healthy donor. Two precipitation methods, acetone precipitation and TCA precipitation, and three digest conditions, i.e. 37°C for 15 minutes or 4 hours, 50°C for 15 minutes and the addition of CaCl2 were assessed [14]. This resulted in eight experimental conditions with five replicates. All samples were processed in parallel and split in separate conditions following the protocol.

For the acetone precipitation, 900µL of ice-cold acetone (-20°C) was added to the samples, followed by centrifugation for 10 minutes (16.000g, -10°C). Next, supernatant was removed, and the remaining pellet was kept at room temperature until dry. The pellet was resuspended in 50µL of digest solution (20ng/µL Trypsin/LysC (Promega), 1mM CaCl2, 5 %(v/v) Acetonitrile in 500mM TEABC and incubated for 15 minutes or 4 hours at 37°C.

For TCA precipitation, 50% (v/v) TCA was added followed by 10 minutes incubation at 4°C and 10 minutes centrifugation at 16.000g. Supernatant was removed, and the pellet was washed with ice-cold acetone (-20°C). Next, the pellet was dried at room temperature and resuspended in 50µL digest solution, follow by incubation for 15 minutes or 4 hours at 37°C.

##### *Resuspension of samples, sample containers and stability*

1 µg of digested recombinant NCAP_SARS2 or SPIKE_SARS2 proteins was resuspended in 100 µL of 0.1% formic acid (FA) in water. These solutions were further combined and 1/10 diluted with either (1) 0.1% FA in water or (2) 10/90/0.1 ACN/H2O/FA. Note: we hence made a mixture at this point, i.e. the final ACN concentration in (2) is 8%. All dilutions were made in threefold. Dilutions were made in either "spring inserts" (SI) (Micro-inserts spring 0.1 mL, Filter Service S.A, Eupen, Belgium) or QuanRecovery Vials (Waters Corporation, Milford, MA), each dilution was injected in triplicate. The injection volume was 0.5 µL. Dilutions were stored at 10°C in the autosampler for 24 hours and then reinjected.

To further test stability, two days later, out of one of the QuanRecovery Vials, stored in the autosampler, a dilution 1/50 in either (1) or (2) was made. Note, the dilution with (1) originated from a solution in (1), the dilution with (2) from a solution in (2). Again the dilutions were made in threefold and each dilution was injected three times. At this point, 10 µL was injected. Samples were stored for 48 hours, at 10°C in the autosampler.

As the above experiment was done in neat solvent, and matrix components might have an influence on stability, a follow-up experiment was conducted. For this, 2 dilution series in UTM were solved, either with 85 µL of (1) 0.1% FA in water or (3) 5/95/0.1 ACN/H2O/FA. We then transferred half of the solution to a spring insert/half of the solution to a QuanRecovery vial. All samples were injected immediately. The injection volume was 10 µL. Note, because of the latter, dilutions were made in max 5% ACN in order not to disturb the chromatography for the early eluting peaks. After injection, samples were stored at 20°C and reinjected after 24 hours and after 48 hours, while storing them at 20°C. Between each injection series, fresh portions of a benchmark (stored at -20°C) were injected as a system function check (to verify for instrument variability). The benchmark was a 1ng/µL solution of recombinant NCAP_SARS2 or SPIKE_SARS2 proteins of which 0.5 µL was injected.

##### *Selection of the UPLC column*

Six different columns were tested, in series using the exact same solvents and gradient. Three types of samples were injected: Blanc, a mixture of the digested recombinant proteins and a pool of 5 digested patient samples with a high viral load. The recombinant proteins were measured first with an open time window in order to determine the retention time. Once the detection window was narrowed down and optimized for each single target peptide on each individual column, the recombinant proteins and the pool of samples were injected twice. Comparison between columns was based on the TargetLynx (Waters, Milford, UK) outcome for area under the curve (sum of all transitions monitored) and S/N for the patient pool. Results are expressed relative compared to the Acquity UPLC Peptide BEH C18 1.7 um column. None of the columns stood out in positive way. We opted for the 300A version, because of long term robustness. The following columns were kindly supplied by Waters Corporation for evaluation: Acquity Cortecs UPLC C18+ 1.6um (P/N:186007114), Acquity UPLC BEH C18 1.7 um (P/N: 186002350), Acquity UPLC Peptide BEH C18 1.7 um (P/N: 186003554), Acquity UPLC Peptide BEH C18 1.7 um 300A (P/N: 186003685), Acquity UPLC Peptide CSH C18 1.7 um (P/N: 186006936), Acquity UPLC Peptide HSS T3 1.8 um (P/N: 186008754).

##### *Incorporating solid-phase extraction (SPE) to increase sample loading*

The application of Solid-phase extraction (SPE) was evaluated using the Waters Oasis Mixed-Mode Cation Exchange (MCX) µElution 96-well plate and spiked eSwab medium (NCAP: 0.16ng/µL and SPIKE: 0.1ng/µL). An experiment design was set-up to assess the following three protocols: (i) 50µl of medium precipitated and digested in 50µl, which was next supplemented with 5µl of 11% FA in a sample vial, (ii) five times 50µl precipitated, each digested in 10µl and merged back into 50µl, which was next supplemented with 5µl of 11% FA and (iii) the same as in (ii) but after digestion SPE was performed. Each protocol was performed in quintuplicate, to assess the assay reproducibility. SPE was performed by diluting the protein digestion supernatant 1:1 with a 4% phosphoric acid solution in water, to quench trypsin activity. The Oasis µElution MCX plate was conditioned and equilibrated by drawing through 200 µL of respectively, Methanol and Water. Five individual acidified protein digests (5*50µL sample) were loaded on one single cartridge, followed by a washing step with 200 µL of 2% FA in water and 200µL of 5% methanol in water. Elution was performed in a QuanRecovery 96 well-plate using 2x25µL of a $NH_40H$ solution in 60/40 $H_20$/ACN. To each well, 5µL of an 11% FA solution was added to obtain a final concentration of 1%FA in 55µL. Finally, 10 µL of each sample was injected for analysis with the Waters Xevo TQ-S using our in-house optimized MRM method.

#### Data acquisition optimization

Each lab was supplied with digested recombinant protein and a triplicate dilution series in UTM. We also supplied them with a Standard Operating Procedure (SOP) describing the development of the in-house MRM method. The participating labs were invited to optimize the method for their specific instrument setup, still some labs decided to incorporate the method directly without any changes. Detailed descriptions of the acquisition strategies can be found in Supplementary Data 14, as well in the raw data.

#### Data Analysis optimization

To construct a data-driven scoring function that decides whether the MRM data was generated by a sample from a COVID-19 patient or from a healthy individual, a machine learning (ML) model was trained and evaluated. The train and evaluation data consisted of a Skyline document results export of 70 LC-MS experiments accompanied by meta data (healthy or diseased) as determined by RT-PCR (**Supplementary Data 8**). Given the limited dataset size (70 patient samples), we opted for a nested cross validation (CV) scheme in which the inner CV optimizes the hyperparameters by means of a grid search, and the outer CV evaluates the model trained on the best hyperparameters as defined by the inner CV.

The inner CV uses a Leave-One-Out (LOO) approach to maximize the usage of the limited amount of data, while the outer CV uses a standard 10-fold CV. The latter allows us to repeat the procedure with different pseudorandom fold splits. The full CV scheme was repeated three times, resulting in 30 final models being trained and evaluated (3 repeats of the 10-fold CV). The full script is available on Github (www.github.com/compomics/Cov-MS-scoring).

The processed Skyline export resulted in 433 features, including meta data such as type of swab medium, UTM or eSwab, which were encoded as features as well, and others that are on the MRM transition level (height, width, and area of the peak, background signal, retention time deviation), as well as on the precursor level (e.g. summed area for all transitions of the precursor). To reduce the unfavorable ratio of features to the number of samples (433/70), principal component analysis (PCA) was performed prior to the training of the ML model. For the classification task, a linear supported vector machine (SVM) algorithm was selected. These two methods, PCA and SVM, provide two hyperparameters that require optimization in the inner CV grid search, i.e. the number of PCA components and the SVM's C regularization term.

Next, the 30 models were evaluated on their respective test fold by calculating the area under the receiver operating characteristic curve (ROC-AUC). This resulted in a median test ROC-AUC score of 0.9167. Furthermore, by analyzing the contribution of each of the initial features to the final principal components, we can estimate the combined importance of each MRM precursor and transition feature set. This can aid in assessing the diagnostic effectiveness of each of the precursors and transitions within the MRM assay. For instance, based on this (limited) dataset, we can assume that SARS-CoV-2 peptides AYNVTQAFGR and GWIFGTTLDSK provide a higher diagnostic value in an MRM assay than the peptides LNQLESK and KQQTVTLLPAADLDDFSK. This illustrates how data-driven approaches, such as ML, do not only facilitate downstream data analysis, but can also help to improve experimental workflows.

Finally, because the significant impact of the media, we trained the models on eSwab and UTM separately.

#### Towards a clinical MRM assay (Green)

##### A Cov-MS QconCAT construct assess sampling efficiency, sample preparation and data acquisition.

After we had generated a list of 17 target peptides, we reached out to PolyQuant to synthesize a QconCAT construct. The target peptides were concatenated into a synthetic polypeptide. For the NCAP protein, natural flanking sites were included to guarantee similar digestion of the QconCAT to the native protein. At the N-terminus, an expression tag was added to ensure high-level expression in E.coli [15]. At the C-terminus, a histidine tag (His-tag) was added to enable protein purification. Three peptides derived from the LC-MS/MS calibration standard RePLiCal were included to validate the quality of the recombinant protein and to monitor the LC of the assay. Additionally, four histone peptides were added with respect to monitor nasopharyngeal sampling quality. Briefly, a QconCAT construct containing these 24 peptides was designed and produced with minor modifications as described previously [16].

Subsequent quality control by LC-MS/MS in DDA mode showed >99.9 % labelling efficiency and >99 % purity of the 15N labelled QconCAT (data not shown).

#### RT-PCR Accreditation Standards are not easily transposable to MRM assays

##### qPCR assay protocol:

RNA was extracted from 140 µL swab collection medium using the QIAamp Viral RNA Mini Kit (Qiagen, Hilden, Germany) according to the manufacturer's instructions. Elution of RNA was realized in a volume of 60 µL elution buffer. Two singleplex, hydrolysis probe-based RT-qPCR assays targeting the nucleocapsid protein gene were performed. Both assays, herein called N1 and N2, were published by the US CDC. Primer and probe sequences can be found in table 1, along with their final concentration. Besides the primers (Biolegio, Nijmegen, The Netherlands) and probe (Integrated DNA Technologies, Coralville, IA, USA), RT-qPCR reactions contained 1X QScript 1-step RT-qPCR Toughmix (Avantor, Radnor, PA, USA), and 5 µL of eluted RNA. The final reaction volume was adjusted to 20 µL with nuclease free water (supplier). RT-qPCR was performed using a LightCycler 480 II (Roche, Basel, Switzerland). Thermal cycling consisted of 10 min at 50 °C and 3 min at 95 °C, followed by 45 cycles of 30 sec at 95 °C and 30 sec at 55 °C. Determination of the quantification cycle (Cq) was performed by the LightCycler 480 Software.

| Assay | Target gene | Oligonucleotide name | Sequence (5' – 3') | Concentration (nM) |
|---|---|---|---|---|
| N1 | Nucleocapsid (N) | N1 Forward primer | GACCCCAAAATCAGCGAAAT | 200 |
| | | N1 Reverse primer | TCTGGTTACTGCCAGTTGAATCTG | 200 |
| | | N1 Probe | FAM-ACCCCGCATTACGTTTGGTGGACC-ZEN/Iowa Black | 200 |
| N2 | Nucleocapsid (N) | N2 Forward primer | TTACAAACATTGGCCGCAAA | 200 |
| | | N2 Reverse primer | GCGCGACATTCCGAAGAA | 200 |
| | | N2 Probe | FAM-ACAATTTGCCCCCAGCGCTTCAG- ZEN/Iowa Black | 200 |

##### *NRC standard*

The Belgian National Research Council (NRC) provided us with their standard of purified inactivated virus. We made a 1/2 dilution series for both UTM and eSwab media spiked with inactivated virus. Both dilution series were measured with our in-house RT-PCR and MRM assay.
Both the N1 and E gene show a near-linear correlation over the dilution series between a Ct value of 22 and 38.For LC-MS purposes, 250 µL of each dilution (20 in total) was spiked with 3.125 ng QconCAT before the samples were split in 5x50µL. Acetone precipitation was performed through the addition of 7 volumes ice-cold acetone (350µL), followed by a centrifugation step at 16000 rpm for 10minutes at 0°C. The supernatant was removed, and the pellet was left to dry at room temperature. 50µL of a 0.01 µg/µL Trypsin-Lys C solution in 50 mM TEABC and 5% ACN was added and the samples were incubated for 15 minutes at 37°C. Finally, SPE was applied in a similar way as described earlier. Note, elution was performed in a QuanRecovery plate using 25 µL of a NH40H solution in 60/40 H20/ACN. Finally, the samples were acquired with a Waters Xevo TQ-XS using two transitions for each target peptide. During data analysis in Skyline it was noticed that the UTM dilution series saturated the column and caused carry-over in the eSwab dilution series (data not shown).

##### *Dilution series with diagnosed negative patients (**supplementary data 19**)*

A dilution series of four Covid-19 cases, confirmed by the in-house qPCR, was created to mimic the NRC standard dilution experiment. These four patient samples were specifically chosen for their low Ct-values (15-20) and because of the different storage media being eSwab, UTM, Virocult and Bioer. 50 µL of each dilution was processed in a similar way as described for the NRC standard and finally SPE was performed with elution in 12,5 µL $NH_40H$ solution in 60/40 $H_20$/ACN, which was further diluted with 7.5µL of a 2% FA solution in water. Note the different sampling volumes between the NRC and the patient dilution (250 vs. 50µL) and the absence of QconCat spike in. The samples were acquired as described for the NRC standard.

### References


[1] P. Navarro, J. Kuharev, L. C. Gillet, O. M. Bernhardt, B. MacLean, H. L. Röst, S. A. Tate, C.-C. Tsou, L. Reiter, U. Distler, G. Rosenberger, Y. Perez-Riverol, A. I. Nesvizhskii, R. Aebersold, S. Tenzer, *Nat. Biotechnol.* **2016**, *34*, 1130.

[2] B. C. Searle, L. K. Pino, J. D. Egertson, Y. S. Ting, R. T. Lawrence, B. X. MacLean, J. Villén, M. J. MacCoss, *Nat. Commun.* **2018**, *9*, 5128.

[3] S. Kim, P. A. Pevzner, *Nat. Commun.* **2014**, *5*, 1.

[4] H. Barsnes, M. Vaudel, *J. Proteome Res.* **2018**, *17*, 2552.

[5] M. Vaudel, J. M. Burkhart, R. P. Zahedi, E. Oveland, F. S. Berven, A. Sickmann, L. Martens, H. Barsnes, *Nat Biotechnol* **2015**, *33*, 22.

[6] B. Mesuere, B. Devreese, G. Debyser, M. Aerts, P. Vandamme, P. Dawyndt, *J. Proteome Res.* **2012**, *11*, 5773.

[7] T. Van Den Bossche, P. Verschaffelt, K. Schallert, H. Barsnes, P. Dawyndt, D. Benndorf, B. Y. Renard, B. Mesuere, L. Martens, T. Muth, *J. Proteome Res.* **2020**, *19*, 3562.

[8] R. Gabriels, L. Martens, S. Degroeve, *Nucleic Acids Res.* **2019**, *47*, W295.

[9] R. Bouwmeester, R. Gabriels, N. Hulstaert, L. Martens, S. Degroeve, ‡ †vib-Ugent, *bioRxiv* **2020**, 2020.03.28.013003.

[10] P. Verschaffelt, P. Van Thienen, T. Van Den Bossche, F. Van der Jeugt, C. De Tender, L. Martens, P. Dawyndt, B. Mesuere, *Bioinformatics* **2020**, *36*, 4220.

[11] H. M. Berman, J. Westbrook, Z. Feng, G. Gilliland, T. N. Bhat, H. Weissig, I. N. Shindyalov, P. E. Bourne, *Nucleic Acids Res.* **2000**, *28*, 235.

[12] T. Vermeire, S. Vermaere, B. Schepens, X. Saelens, S. Van Gucht, L. Martens, E. Vandermarliere, *Proteomics* **2015**, *15*, 1448.

[13] "PyMOL | pymol.org," can be found under https://pymol.org/2/#products, **n.d.**

[14] P. Glibert, K. Van Steendam, M. Dhaenens, D. Deforce, *Proteomics* **2014**, *14*, 680.

[15] Z. M. Al-Majdoub, K. M. Carroll, S. J. Gaskell, J. Barber, *J. Proteome Res.* **2014**, *13*, 1211.

[16] J. M. Pratt, D. M. Simpson, M. K. Doherty, J. Rivers, S. J. Gaskell, R. J. Beynon, *Nat. Protoc.* **2006**, DOI 10.1038/nprot.2006.129.
